# Ergonomic Interventions and Technology in Preventing Musculoskeletal Disorders Among Sonographers (2000-2025): A Systematic Mapping Review

**DOI:** 10.64898/2026.09.15.26363110

**Authors:** Ama Boahene Akomah, Albert Dayor Piersson, Marizuk Waris Tizumah

## Abstract

The sonography profession faces a persistent occupational health crisis, with nearly 90% of practitioners reporting work-related musculoskeletal disorders (WRMSDs) despite two decades of ergonomic education. This study performs a definitive bibliometric analysis to map the evolution of ergonomic technology and interventions for WRMSDs from 2000 to 2025. Addressing this unabated prevalence of injury is paramount to ensuring the long-term sustainability and resilience of the imaging workforce amid rising clinical demands. Adhering to PRISMA guidelines, the methodology utilized the Scopus database to extract a curated dataset of 93 peer-reviewed publications. Quantitative and network analyses were performed using VOSviewer and the Bibliometrix R-package to evaluate publication growth, leading contributors, and thematic clusters. The findings reveal a conceptual shift from foundational hazard identification toward high-order technological solutions, such as workstation automation and specialised transducer handles. Notably, the results indicate that traditional education-only models have reached a maximum threshold, as environmental constraints and high patient volumes frequently override individual behavioural changes. By providing the first integrated evaluation of the transition from awareness to technology-driven intervention, this study fills a critical gap in the literature regarding the structural role of technology as a prevention support tool. In practice, these findings provide a roadmap for hospital administrators and manufacturers to prioritise proactive environmental transformation over individual posture correction to safeguard practitioners’ health.

## 1. INTRODUCTION

Diagnostic medical sonography as a profession is one of the key pillars of contemporary healthcare provision, offering essential real-time imaging, non-invasive diagnostic solutions, and radiation-free interventional guidance that can provide global clinical stability [1–4]. Currently, the industry has a much broader impact in specialities including obstetrics, cardiology, and musculoskeletal medicine [2,5–7]. However, advanced technology, high patient workload and the ageing population have generated heightened imaging pressure, compelling the sonography owners of business and hospital administration to make trade-offs in operational throughput versus practitioner health [8–12].

Work-related musculoskeletal disorders (WRMSDs) have continued to be on the list of the most important causes of occupational injury in the industry, with high prevalence rates [8,12–15]. Pain associated with the scanning process is reported by approximately 90 percent of diagnostic medical sonographers and vascular technologists, which has been occurring historically and has not substantially changed in 20 years [7,8,12,14,16]. The sustainability of the operations approaches in the industry has become a topic of global concern [15,17,18]. The old paradigm of high-volume/low-rest models is still the predominant mode of operation in the world, with its burden of biomechanical loads and inability to address the social and physical aspects of sonographers’ well-being [1,9,16,19]. It has prompted the increase of stakeholder attention to the issue of ergonomic compliance and the enhancement of the workplace safety requirements [5,12,15, 21].

Health and safety requirements are supported by policy frameworks and professional guidelines to cope with this crisis [5,14,15,18]. To balance the diagnostic effectiveness, workability of the equipment, and the safety of the practitioners, medical imaging departments must embrace ergonomic interventions [7,9,17,20]. These targets presuppose a swift and massive change of the current awareness-based education to the proactive and intervention-based approaches [10,12,21–23]. These biomechanical pillars are now also highly valued by the industry globally, and social sustainability is also being introduced with better workstations and mental assistance [1, 6,17,19]. However, the infrequent quality of rests and the sophistication of the current patient demographic, i.e., the increase in obesity, present technical and operational uncertainties that the conventional ergonomic training is not always able to define [10,13,20,24]. Risk management has to deal with integration issues because the number of portable examinations demanded and the fragmentation of clinical workflows grow [3,10,16,19]. Although there has been an extensive adoption of ergonomic principles, there are gaps in implementation, as a majority of practitioners find it difficult to quantify the intersection between workstation adjustability and real musculoskeletal relief [7,12,18,22]. Such limitations imply that the available behavioural models do not suit the present times when technological innovation forms a mandatory condition to survive in the profession [7,15,17,25].

Sonography has a lot of technological assets, which include adjustable examination beds and height-adjustable PACS workstations that maximise scanning positions [5,9,15,16,20]. In addition, microbreak software and surface Electromyography (EMG) biofeedback may also be useful in supplementing real-time posture monitoring and preventing cumulative trauma [7,18,22]. Multiple media training and digital tutorials facilitate ergonomic literacy at the standard level [21–23]. Such efforts are in line with the priorities of the sector to clinical excellence and practitioner safety and reveal the interrelatedness of technological innovation and injury prevention [7,15, 17,18].

Moreover, the inclusion of these technologies provides an avenue to the establishment of resilience among sonography practitioners. Investigations approximate that scaling workstation automation and the ability to use both hands could substantially decrease shoulder abduction, decrease reach, and avoid long-term nerve entrapment [3,7,13,24]. The institutional framework behind this change is strong, as there is more cooperation between manufacturers, professional societies, and ergonomists to create safer ecosystems [8,15,17,18]. In spite of these positive circumstances, the complete adoption of the technology is not widespread in reality [10,12,22]. The obstacles are expensive equipment, resistance of the administration, and the inability to overcome the established scanning habits [9,10, 19,21].

Nonetheless, technological implementation by the sector has become very advanced by undertaking various global projects. The most prominent organisations have tested participatory ergonomic systems and put in place design interventions, which showed the gains of pain reduction, which can be used alongside clinical throughput [7,17,18]. In addition to general imaging usage, technology has also been used in contexts that are more wide-ranging, including specialised obstetric platforms and ultrasound-guided tools of interventional pain management [2,4,5]. In the same manner, neuro-muscular surveillance plans are under trial to detect fatigue before injury [1,7,13]. All these practical applications demonstrate that the technological dimension of the contemporary sonography model of safety is a crucial aspect [7,14,15,17,18].

Considering these opportunities and challenges, a thorough mapping of the research field is essential. Bibliometric analysis is a strong quantitative research approach that allows assessing research structures, dynamics, and impact [26,27]. The method has already been used to apply to different fields in previous research, including the mapping of occupational hazards and technological development in healthcare [28]. Corresponding initiatives have been placed on regional rates and the efficiency of specialised training [9,21–23].

However, in spite of an increasing amount of literature, the exact shift between ergonomic awareness to technology-based intervention that is being examined in the research regarding sonographers has not been evaluated in a more definite and thorough manner. Rather, current studies have concentrated on both singular prevalence rates and broad postural instructions, giving minimal information about the structural role of technology as a preventive factor [8,14,18]. This inability to create an integrated, technology-driven mapping constrains the capacity of policymakers and hospital managers to develop specific interventions in respect of the new imaging reality [12,15,17]. This study helps in filling gaps in knowledge and offering practical information on researching the topic of fair Internet access by systematizing the growth in publications, most prolific authors, and thematic areas through a final dataset comprising 93 publications reviewed.

### 1.1 Review and Aim

The study aimed to examine the current trends in the development of ergonomic technology and intervention of WRMSDs. To do so, the study will answer the following review questions (RQ):

(RQ1) What is the growth trend of studies on sonographer ergonomics and WRMSDs in the globally?

(RQ2) Which authors, journals, and countries are more likely to influence the ergonomic landscape, and how do they work together?

(RQ3) What are the thematic clusters of research regarding sonographer sustainability, especially regarding biomechanical, environmental, and technological aspects?

(RQ4) What are the future trends and changing research interests of sonographer safety, particularly in relation to workstation automation and participatory ergonomics? and

(RQ5) What do the available gaps in the literature mean as regards developing a comprehensive technology-based prevention model for sonographers?

To address these, this research uses bibliometric tools to offer a general evaluation of the subject. The findings will be beneficial because:

(i) the researchers will discover the opportunity to cooperate

(ii) the practitioners will achieve the benchmark of technology integration, and

(iii) the policymakers will find the alignment of safety frameworks with clinical reality.

The remainder of this paper is structured as follows: Section 2 details the methodological framework, including data sources and metadata quality. Sections 3and 4 presents the Results and Discussion. Section 5 concludes with key findings and directions for future research.

## 2. METHODOLOGY

### 2.1 Research Method

This study follows a systematic and bibliometric protocol consistent with the Preferred Reporting Items for Systematic Reviews and Meta-Analyses (PRISMA) guidelines to ensure transparency, reproducibility, and methodological rigour [29]. The methodological framework used to achieve the objectives of this review is presented in Figure 1. To document the identification, screening, eligibility, and final inclusion of studies on ergonomic interventions in sonography, a PRISMA flow diagram was used as the primary reporting tool.

**Figure 1:**
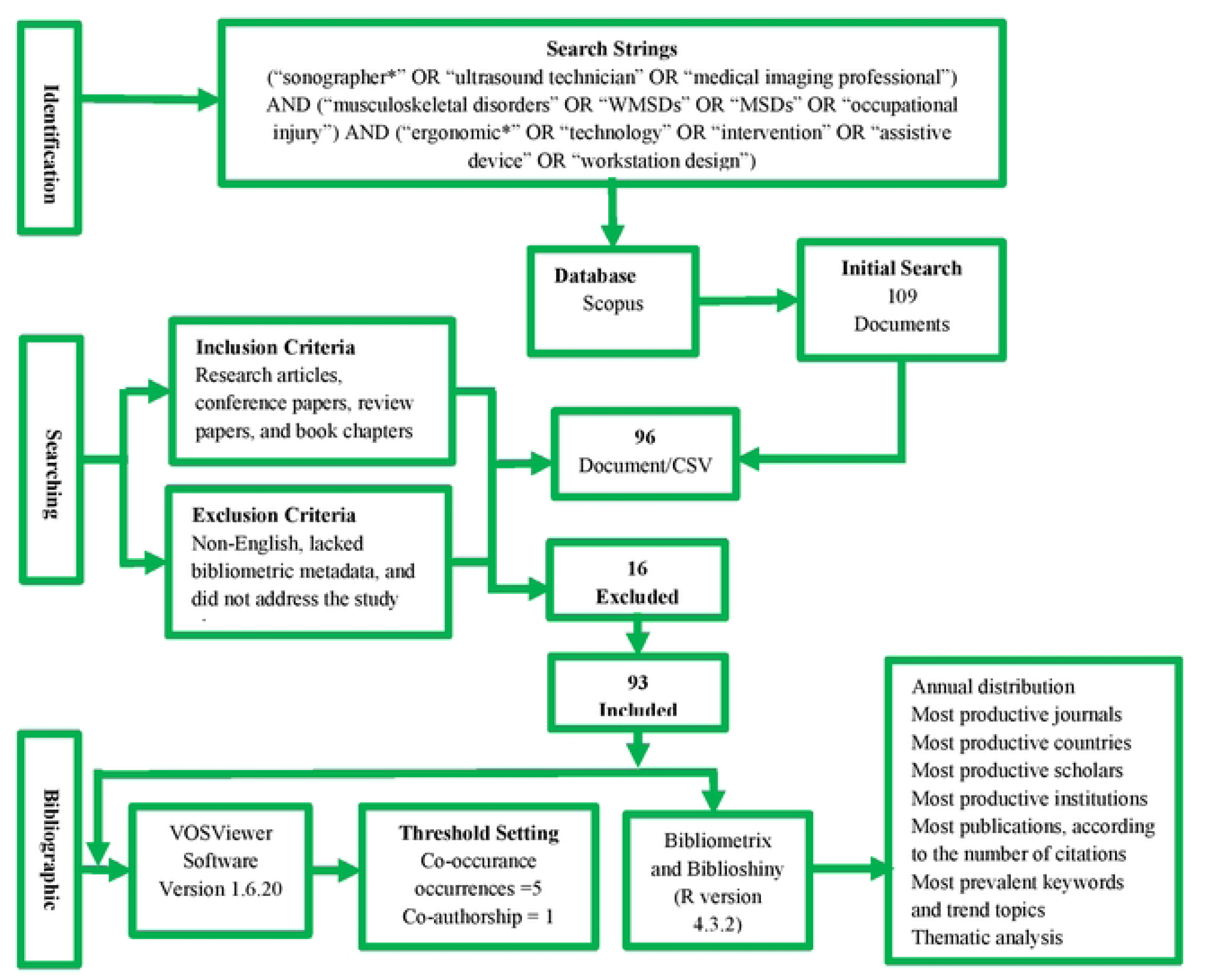
The Overarching Flow Diagram of the Methodology.

### 2.2 Data Source and Search Strategy

Scopus was chosen because it has a wide range of peer-reviewed publications with structured metadata and high indexing levels that facilitate bibliometric studies in the fields of healthcare, ergonomics, and occupational health [28]. Scopus has better reliable bibliographic data, which can be used in systematic and bibliometric studies than Web of Science (only 54% of Scopus records are indexed) and Google Scholar (non-peer-reviewed information not included and lacking organized metadata) [30]. Metadata such as abstracts, keywords, citations, affiliations, and funding information, which are necessary to conduct advanced network analysis, are also available on the platform. The search query was through the Title, Abstract, and Keywords search fields, and a combination of words representing sonography, ergonomics, and musculoskeletal disorders. In order to replicate, the exact search string is as shown below:

> *(“sonographer*” OR “ultrasound technician” OR “medical imaging professional”) AND (“musculoskeletal disorders” OR “WMSDs” OR “MSDs” OR “occupational injury”) AND (“ergonomic*” OR “technology” OR “intervention” OR “assistive device” OR “workstation design”)*

The publication window was set from 2000 to 2025, aligning with the introduction of advanced technological scanning interfaces and the evolving digitalisation of medical imaging systems.

### 2.3 Inclusion and Exclusion Criteria

Peer-reviewed journal articles, conference papers, review articles, and book chapters were included; the publications had to be written in English; and the studies had to cover the topics of ergonomic interventions, technological solutions, or musculoskeletal health among sonographers. Moreover, the subsequent exclusion criteria were used: editorials, notes, letters, commentaries, and errata; articles without abstracts or lacking bibliometric metadata; and articles that were not related to the ergonomic health of practitioners or studied patient clinical outcomes. Two independent reviewers decided all the screening choices, after which any disagreements were settled by discussions to promote the integrity of the final dataset.

### 2.4 Bibliometric and Systematic Analysis

Bibliometric analysis was performed with the VOSviewer version 1.6.20 to co-occurrence of keywords and bibliographic coupling, and Bibliometrix and Biblioshiny (R version 4.3.2) to descriptive performance measures, collaboration networks, thematic maps, trend topics, and factorial analysis [27,31]. In the co-word analysis, two was set as the minimum occurrence of the keywords. In bibliographic coupling, two shared references between the documents had to be present. These cutoffs represent commonplace health sciences and ergonomics bibliometrics to balance their depth of analysis and cluster stability [26,32].

### 2.5 Cluster Validation and Reliability Procedures

The cluster names came up in a two-step validation process. First, the association-strength algorithm of VOSviewer resulted in the creation of clusters that are formed due to keywords and citation relationships [31]. Second, each cluster was reviewed independently by an expert panel of three scholars in the area of ergonomics, occupational health and medical imaging and labelled accordingly. The labelling differences were resolved by consensus. This method enhanced construct validity and was consistent with the procedures that were established in systematic and bibliometric reviews [32,33].

### 2.6 Reproducibility and Data Availability

To support the advancement of ergonomic research, the full dataset, including the exported Scopus CSV file, the search string, the PRISMA flow diagram, and the R analytical scripts, will be made available upon reasonable request.

### 2.7 Data Quality and Completeness Assessment

This section evaluates the integrity of the retrieved bibliographic data as shown in Table 1. Ensuring metadata completeness is a prerequisite for reducing bias in performance analysis and science mapping. The assessment, conducted via automated metadata analysis in the Bibliometrix R package, utilising evaluative nomenclature to categorise the reliability of various bibliographic attributes [27]

**Table 1:** Completeness of metadata of 93 documents from Scopus.

| Metadata | Description | Missing Counts | Missing % | Status |
| --- | --- | --- | --- | --- |
| AB | Abstract | 0 | 0.00 | Excellent |
| C1 | Affiliation | 0 | 0.00 | Excellent |
| AU | Author | 0 | 0.00 | Excellent |

| Metadata | Description | Missing Counts |  | Missing % | Status |
| --- | --- | --- | --- | --- | --- |
| DT | Document Type | 0 |  | 0.00 | Excellent |
| SO | Journal | 0 |  | 0.00 | Excellent |
| LA | Language | 0 |  | 0.00 | Excellent |
| PY | Publication Year | 0 |  | 0.00 | Excellent |
| TI | Title | 0 |  | 0.00 | Excellent |
| TC | Total Citation | 0 |  | 0.00 | Excellent |
| CR | Cited References | 1 |  | 1.08 | Good |
| DI | DOI | 5 |  | 5.38 | Good |
| DE | Keywords | 7 |  | 7.53 | Good |
| ID | Keywords Plus | 10 |  | 10.75 | Acceptable |
| RP | Corresponding<br>Author | 20 |  | 21.51 | Poor |
| WC | Science<br>Categories | 93 |  | 100.00 | Completely<br>missing |

In the first examination of the dataset, a very high level of quality in such fundamentals of metadata is observed. Core attributes important to the performance metrics and descriptive bibliometrics, such as Abstracts (AB), Authors (AU), Titles (TI), Publication Years (PY), and Total Citations (TC), have a 0% missing rate, which is rated as an excellent value. Their fullness is crucial; it means that the following analysis of citation and mapping of the trends according to time is conducted on the full representation of the chosen literature without the possibility of undercounting and misattribution. Moreover, the ideal storage of Affiliation (C1), Document Type (DT), Journal (SO), and Language (LA) data offers a sound system for determining the so-called intellectual pillars and geographical sources of sonographer ergonomics research without being cluttered by data.

In fields related to digital indexing and thematic categorisation, a slight decrease in metadata density is evident. Good References cited (CR), Digital Object Identifiers (DI), and Keywords (DE), and missingness 1.08 to 7.53. Although these non-significant gaps do not pose a statistical threat to a general thematic mapping, they indicate the slight inconsistency in the process of indexing older or regional publications in the main database. Although the status of Keywords Plus can be described as Acceptable (10.75% missing), it also indicates that the data set is good in terms of author-defined themes. However, the computer-generated thematic indexing is rather less comprehensive. This inconsistency leads to the need to place more trust in Author Keywords and Abstract analysis so as to provide a holistic thematic synthesis, especially when cross-comparing the emerging ergonomic interventions.

The greatest constraints in the dataset are related to administrative and disciplinary metadata. The field with the Corresponding Author (RP) classification is classified as Poor, and the rate of missing is 21.51%. This reveals a limitation in the application of the granular collaboration analysis by the primary institutional responsibility, but the status of the general Affiliation data as excellent tends to overcome this constraint, since it allows mapping at the institutional level. Most importantly, Science Categories (WC) are found to be missing (100%). One may assume that such a total absence is caused by the limitations in database export or by the non-standardisation of disciplinary tagging of the particular niche of sonography in the source index. This is an indication that the study will have to make use of Keyword co-occurrence and Journal titles (SO) as the means of defining the multidisciplinary core of the field, including not only ergonomics and medical imaging, but also occupational health, without necessarily relying on pre-defined category filters. Regardless of those particular defects, the general score of 11 out of 15 metadata fields, which can be described as the presence of an excellent to a good level, can confirm that the dataset has the empirical rigour required to make a conclusive bibliometric review.

## 3. RESULTS

### 3.1 Data Characteristics and General Overview

The process of systematic retrieval and subsequent filtering (shown in Figure 2) resulted in a final dataset of 93 documents published between 2003 and 2025, distributed among 44 different sources, including high-impact journals, books, and conference proceedings. The dataset recorded a constant average annual growth rate of 8.49%. The mean age of the documents in the collection is 8.92 years, and the documents have an average of 14.15 citations each.

**Figure 2:**
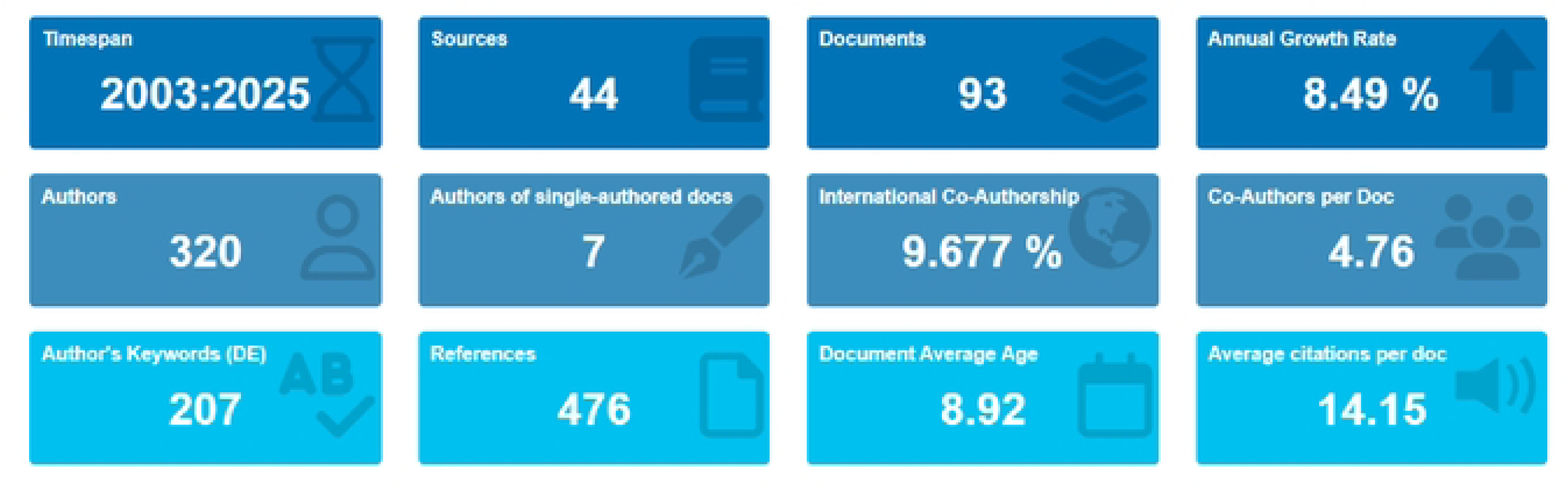
Main Information.

The structure of the document types shows that the largest portion of the dataset is made up of original articles (n = 69), followed by conference papers (n = 12), reviews (n = 9), and book chapters (n = 3). The dataset contains 859 Keywords Plus (ID) and 207 Author Keywords (DE).

On authorship and collaboration, 320 authors contributed to the 93 documents, giving a co-author ratio of 4.76 per document. The number of single-authored documents was 9, and 7 authors published as sole authors. The international co-authorship rate was 9.68%.

### 3.2 Trend in Annual Article Production

The temporal evolution of the literature from 2003 to 2025 reveals a non-linear but upward trajectory, characterised by a compound annual growth rate (CAGR) of 8.49%. As illustrated by the production data in Figure 3, the research landscape can be categorised into three distinct phases: the foundational period (2003 to 2012), the peak-intensification period (2013 to 2019), and the recent volatility and resurgence phase (2020 to 2025).

**Figure 3:**
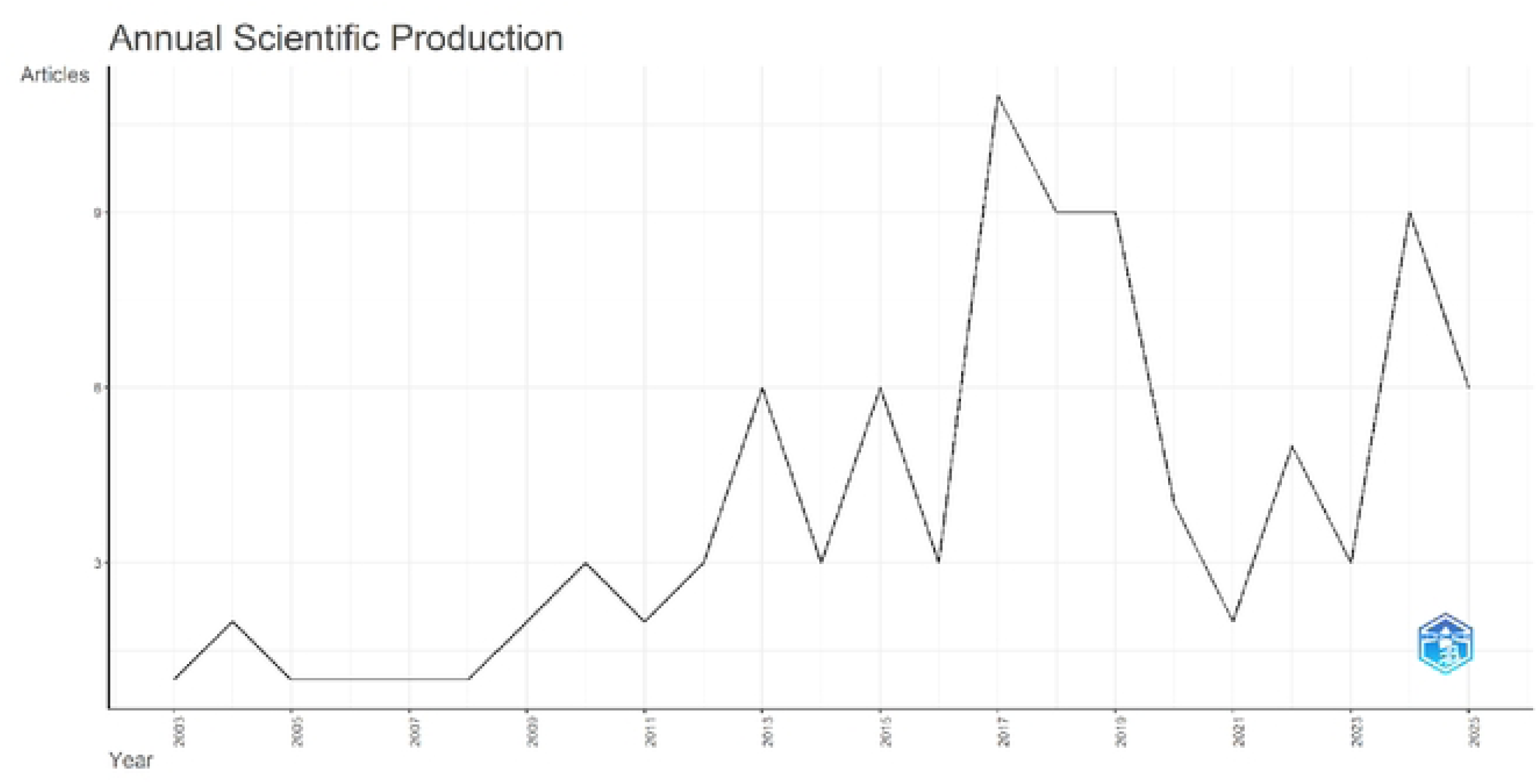
Annual scientific production and growth trend of sonographer ergonomics research (2003–2025)

In the initial years (2003 to 2012), scientific output was infrequent and small-scale, with yearly publications ranging between 1 and 3 articles. Output rose to 6 articles in 2013, a one hundred percent increment over the preceding year. The field was most productive between 2017 and 2019, with a maximum of 11 articles in 2017. A pronounced deviation is observed in the 2020 to 2023 period, where publication volume fell to a minimum of only 2 articles in 2021. The most recent data show 9 articles in 2024 and 6 already indexed in the first part of 2025.

### 3.3 Global Research Collaboration Network

The international co-authorship frequency of 9.68% indicates moderate international integration. Based on the collaboration information in Figure 4, the network is centred on a small number of hubs. The strongest cooperation path is between the USA and China (Frequency = 103.82), matched in strength by the connection between Italy and China (Frequency = 103.82). In the European region, a regional sub-network links Italy and Germany (Frequency = 10.39) and Switzerland (Frequency = 8.21). Inter-regional cooperation is also recorded between Sweden and Bahrain (Frequency = 50.54), the USA and Iran (Frequency = 54.27), and the USA and Saudi Arabia (Frequency = 44.54). Collaboration involving Turkey (Frequency = 35.17) and the Czech Republic (Frequency = 15.31) is also frequent. In contrast, interaction values for countries such as Chile and Mexico are negative or near zero, indicating limited collaborative synergy with the global network.

**Figure 4:**
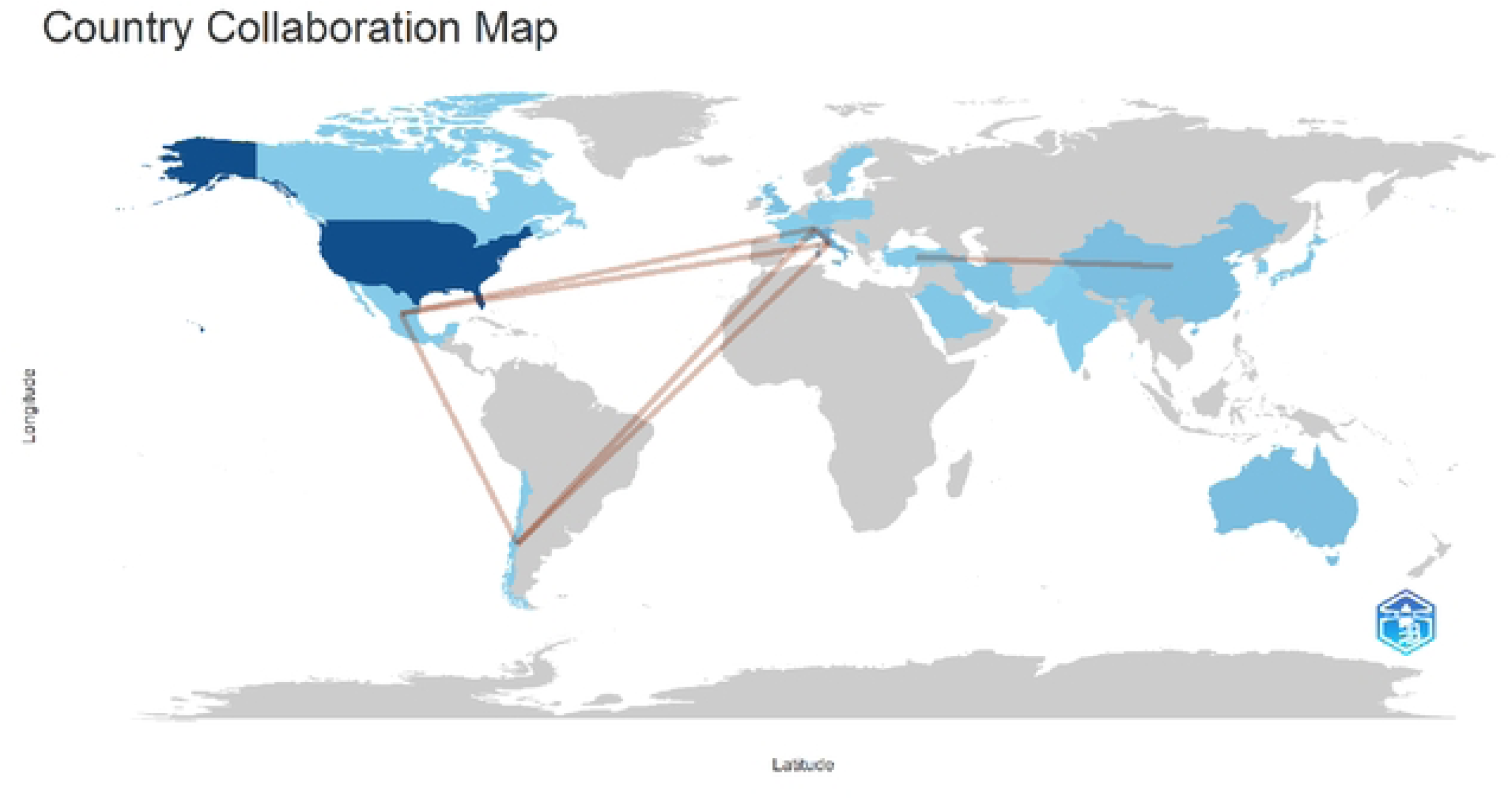
Country collaboration map of sonographer ergonomics research.

### 3.4 Most Occurring Keywords and Trend Topics

As Figure 5 and Table 2 show, the most conspicuous keywords are human (Frequency = 71), ergonomics (Frequency = 70), and article (Frequency = 57). Clinical and technical parameters of the discourse are represented by echography (Frequency = 49), musculoskeletal disease (Frequency = 45), and ultrasonography (Frequency = 30). The demographic keywords female (Frequency = 44), male (Frequency = 41), and adult (Frequency = 39) also feature in the top tier, alongside humans (Frequency = 31).

**Figure 5:**
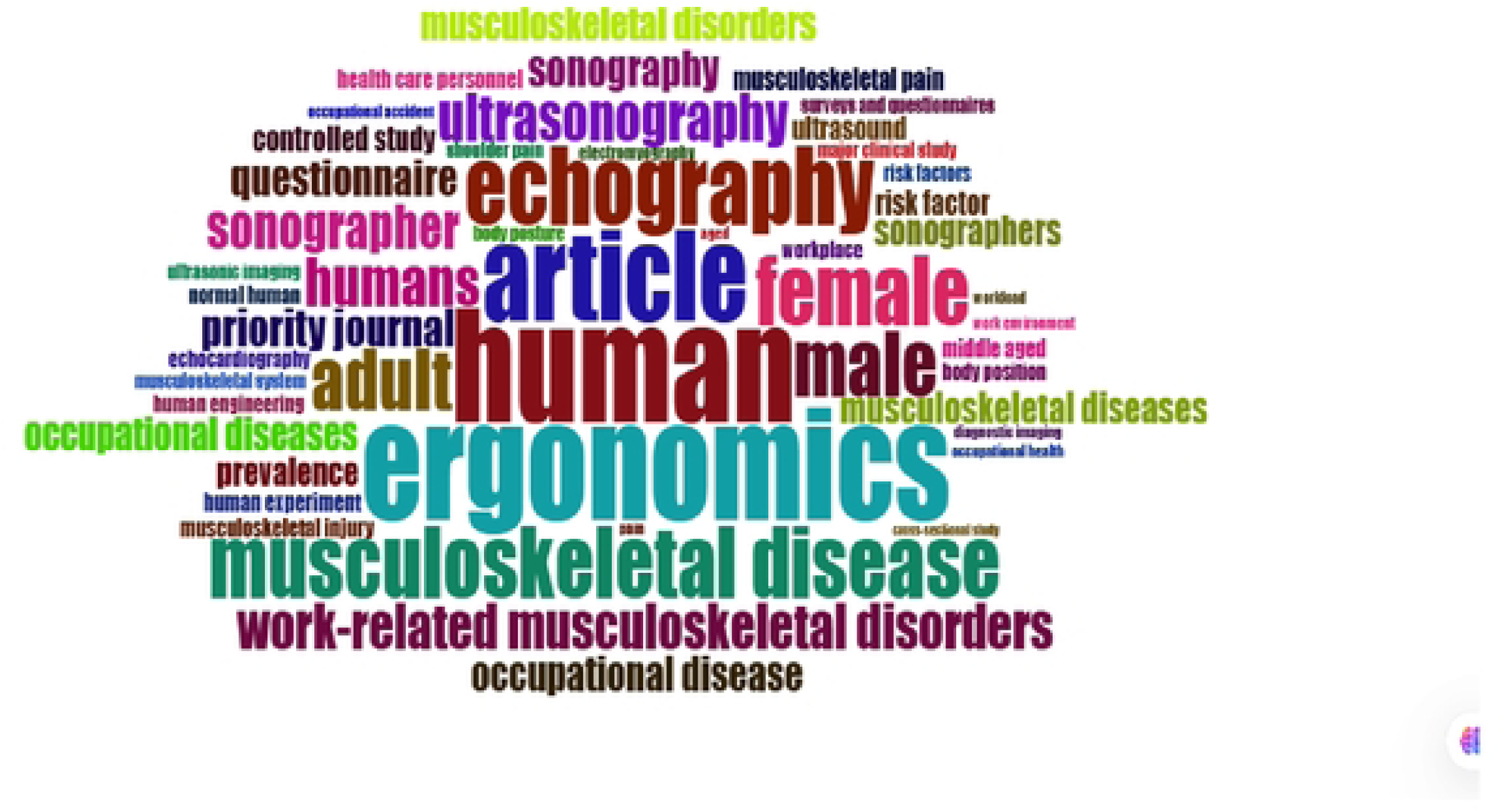
Word cloud of the most frequent keywords in sonographer ergonomics research.

**Table 2:** Top 10 most frequent keywords.

| Rank | Terms | Frequency |
| --- | --- | --- |
| 1 | Human | 71 |
| 2 | Ergonomics | 70 |
| 3 | Article | 57 |
| 4 | Echography | 49 |
| 5 | Musculoskeletal Disease | 45 |
| 6 | Female | 44 |
| 7 | Male | 41 |
| 8 | Adult | 39 |
| 9 | Humans | 31 |
| 10 | Ultrasonography | 30 |

Beyond static word frequencies, the trend topic analysis in Figure 6 tracks the temporal evolution of keywords by median occurrence year. In the early period (2009 to 2013), the most prominent terms were occupational hazard (Median: 2009), bioengineering (Median: 2012), human engineering (Median: 2013), body posture (Median: 2013), and wrist (Median: 2013). In the intermediate period (2014 to 2019), the leading terms were electromyography (Median: 2014), biomechanics (Median: 2015), human experiments (Median: 2016), musculoskeletal diseases (Median: 2016), workload (Median: 2016), female (Median: 2017), and occupational health (Median: 2019). In the current period (2020 to 2025), the most frequent terms were major clinical study (Median: 2020), controlled study (Median: 2021), cross-sectional study (Median: 2021), risk factors (Median: 2021), musculoskeletal pain (Median: 2022), questionnaire (Median: 2022), and work-related musculoskeletal disorders (Median: 2023).

**Figure 6:**
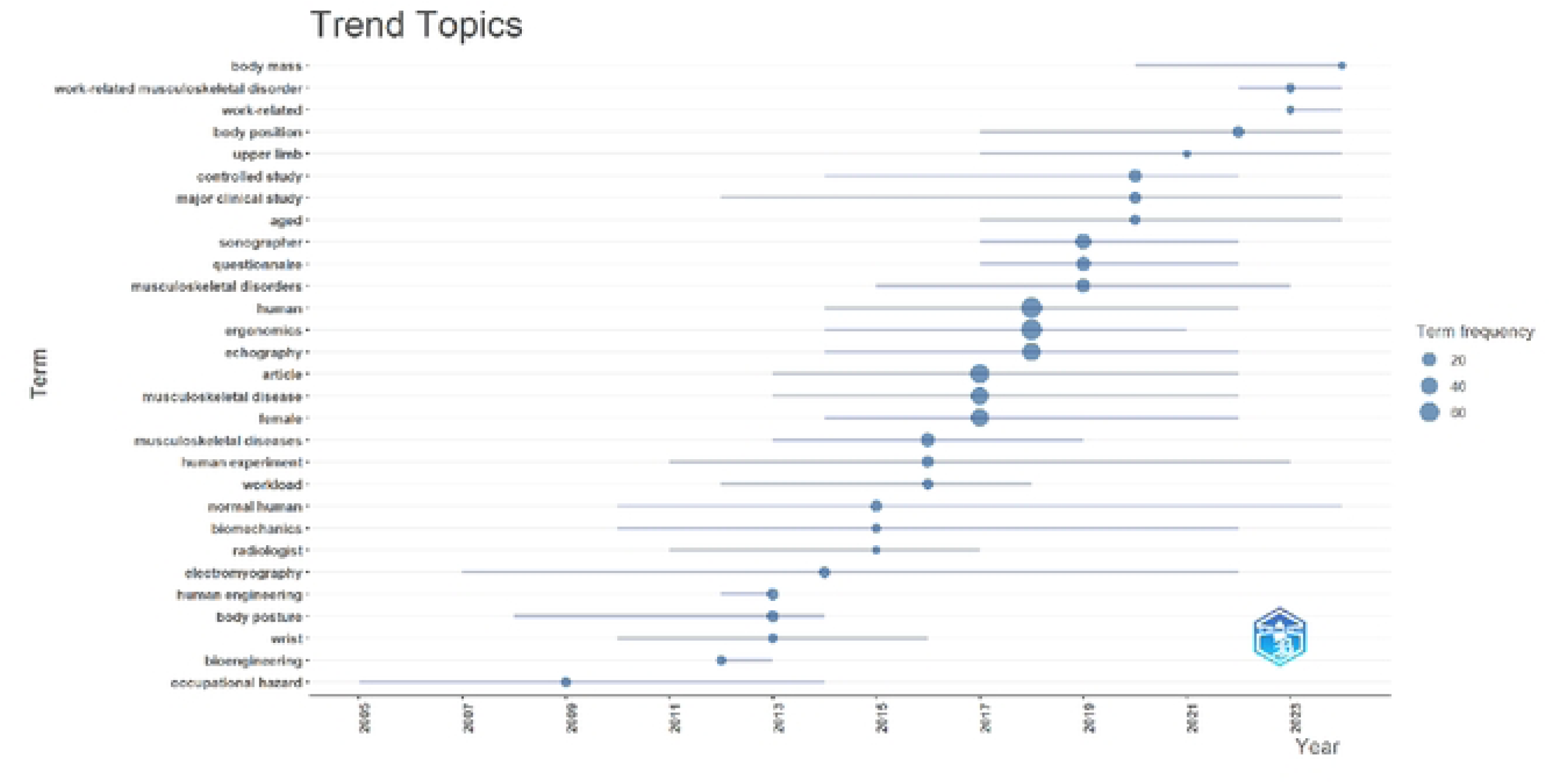
Trend topics in sonographer ergonomics research (2003–2025)

### 3.5 Network Visualisation of Keywords into Clusters

The intellectual structure of the field was outlined using a co-occurrence analysis of author keywords in VOSviewer. With a minimum frequency of 5, 83 keywords were selected from 984 and grouped into three thematic clusters. The network visualisation is provided in Figure 7.

**Figure 7:**
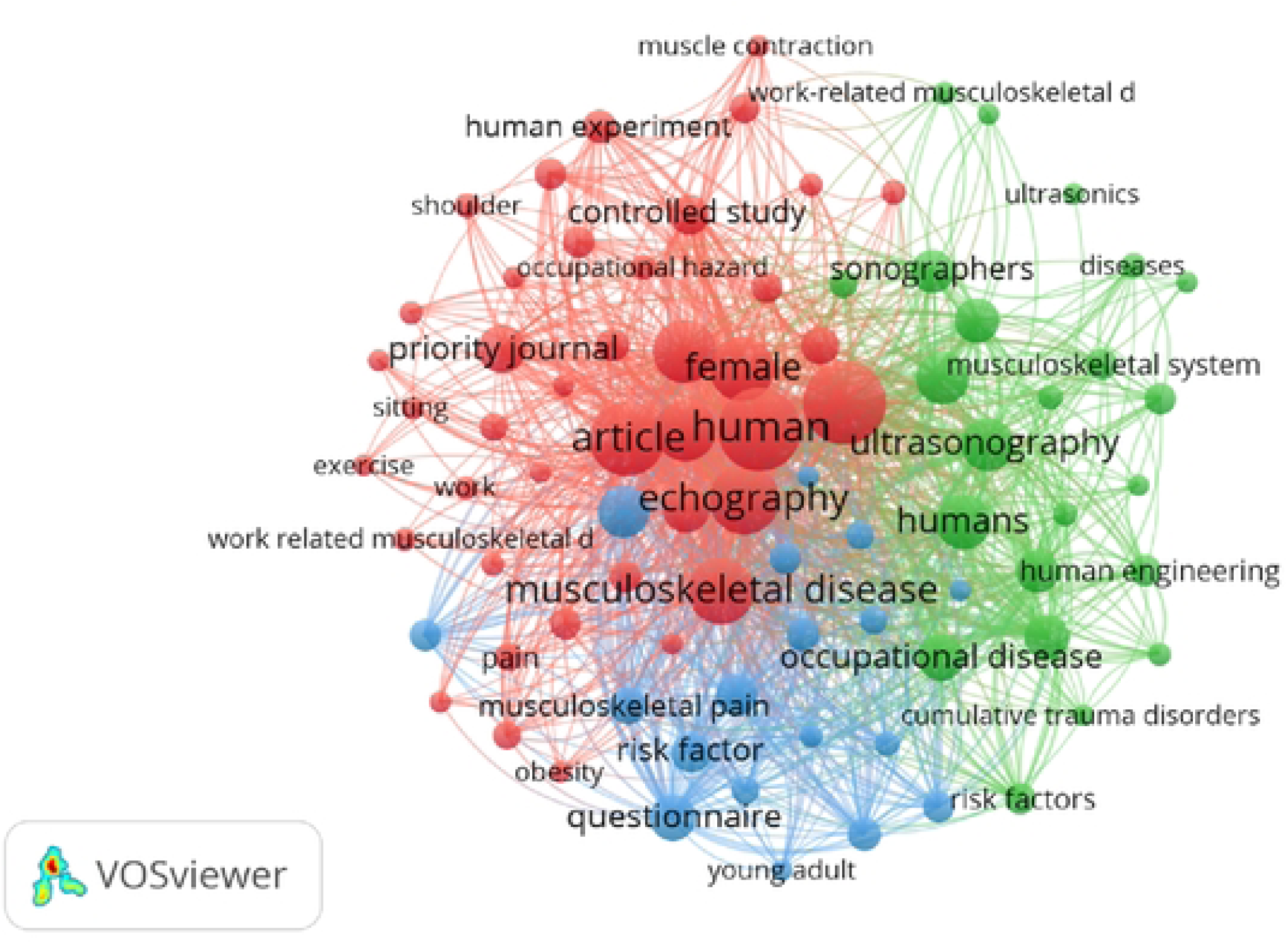
Network visualisation of co-occurring keywords and their Total Link Strength.

Cluster 1 (Red) is the largest cluster (41 items). Its dominant keywords are Ergonomics (TLS = 924), Echography (TLS = 745), Human Experiments (TLS = 176), Body Posture (TLS = 157), Electromyography (TLS = 130), Muscle Contraction (TLS = 82), and Biomechanics (TLS = 74). The cluster also contains the terms Shoulder, Wrist, Review, and Priority Journal.

Cluster 2 (Green) contains 23 items, led by Ultrasonography (TLS = 488), Occupational Diseases (TLS = 398), Work-Related Musculoskeletal Disorders (TLS = 329), Human Engineering (TLS = 156), Occupational Exposure (TLS = 141), and Diagnostic Imaging (TLS = 106). The keyword United States of America also falls within this cluster.

Cluster 3 (Blue) contains 18 items, led by Sonographer (TLS = 437), Questionnaires (TLS = 414), Prevalence (TLS = 349), Musculoskeletal Pain (TLS = 252), Workload (TLS = 158), Cross-sectional Studies (TLS = 147), and Aged (TLS = 154).

### 3.6 Thematic Mapping, Factorial Analysis, and Thematic Evolution of Keywords

The thematic map segments the research landscape into four quadrants. This analysis, as shown in Figure 8, was conducted with a keyword limit of 25 and a minimum cluster frequency of 5. The Motor Themes quadrant (upper right) contains Sonographer, Questionnaire, Prevalence, Musculoskeletal pain, Major clinical study, and Risk factor. The Niche Themes quadrant (upper left) groups Female, Male, and Adult with the technical terms Electromyography, Human experimentation, Body position, and Occupational hazards. The Basic Themes quadrant (lower right) contains Ergonomics, Ultrasonography, Work-related musculoskeletal disorders, Occupational disease, and Human engineering. The lower left quadrant, representing emerging or declining themes, contains Ultrasound, Echography, Diagnostic imaging, Occupational accident, and Musculoskeletal injury.

**Figure 8:**
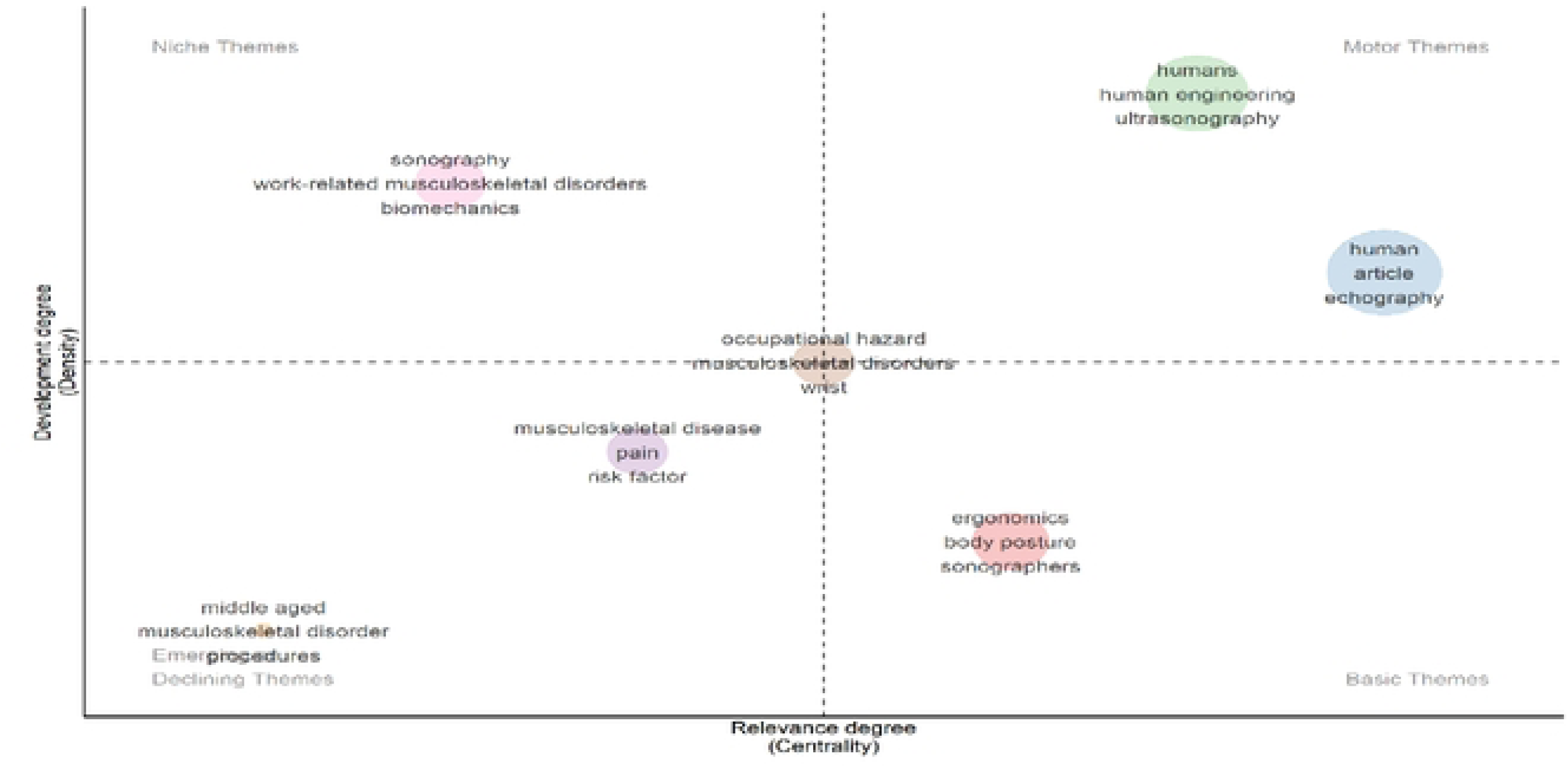
Thematic mapping of keywords into themes.

Figure 9 presents the thematic evolution of the field across four periods (2003 to 2014, 2015 to 2018, 2019 to 2021, and 2022 to 2025), tracked using a Sankey diagram with a weighted analysis (word events) and a minimum weight index of 0.1. In the first period (2003 to 2014), the dominant themes were Ergonomics (n = 47), Sonography (n = 40), Occupational hazard (n = 31), and Musculoskeletal diseases (n = 30). In the second period (2015 to 2018), the leading themes were Echography (n = 99), Humans (n = 93), Work-related musculoskeletal disorders (n = 43), and Awareness (n = 25). In the third period (2019 to 2021), the central keyword Sonographer (n = 23) emerged, together with Prevalence (n = 17) and Exercise (n = 8). In the current period (2022 to 2025), Ergonomics (n = 104), Adult (n = 99), Sonographer (n = 72), and Occupational health (n = 11) reached their highest recorded frequencies.

**Figure 9:**
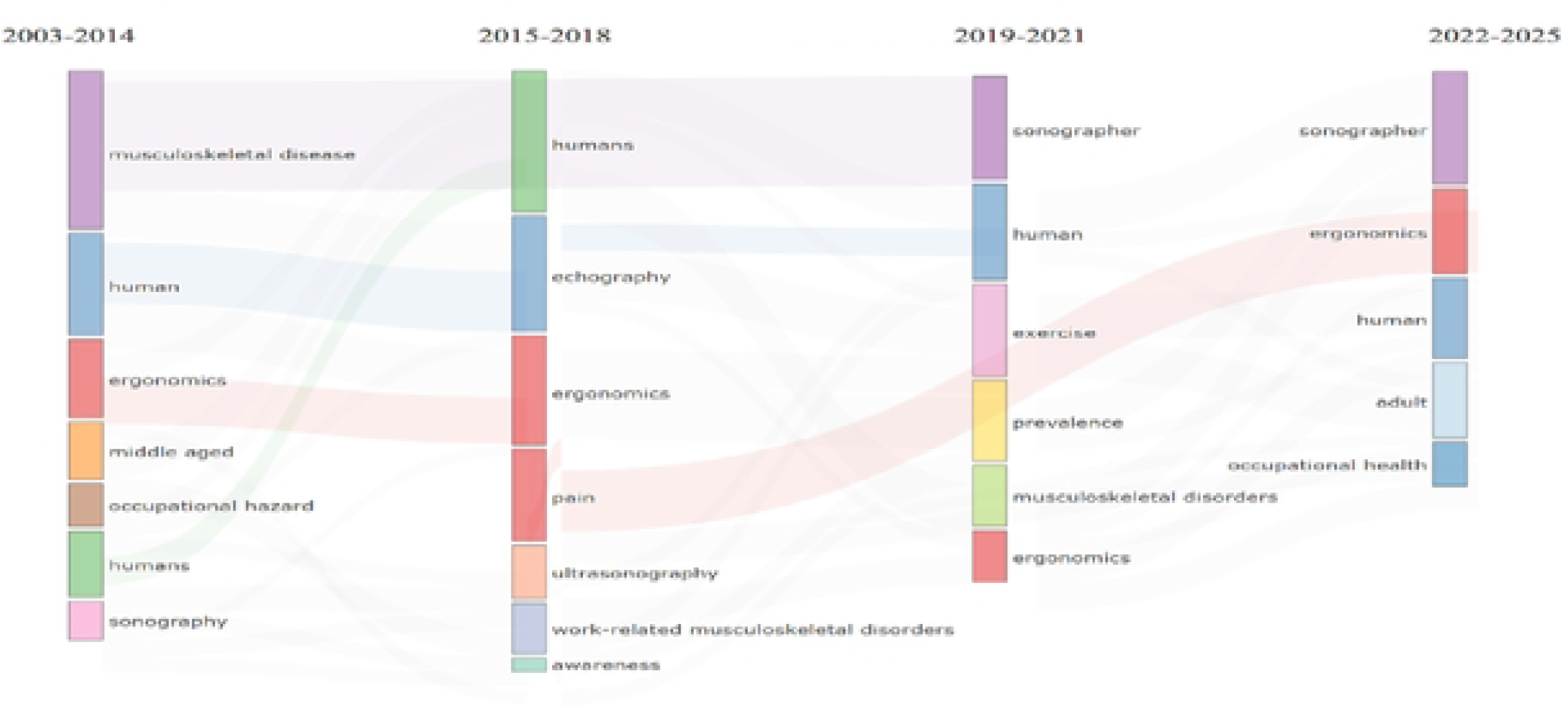
Thematic evolution and the emergence of the sonographer ergonomics.

The conceptual structure of the field was further mapped through Multiple Correspondence Analysis (MCA) of the top 50 keywords, which produced three conceptual clusters in a two-dimensional space, as shown in Figure 10. The largest cluster (Red) combines demographic variables (female, male, adult, aged), clinical modalities (echography, sonography, echocardiography), experimental methods (electromyography, human experiment), and the terms body position, body posture, and workplace. The second cluster (Blue) is marked by prevalence, questionnaire, risk factor, middle-aged, and occupational diseases. The third cluster (Green) contains sonographers and work-related musculoskeletal disorders (WMSDs). The terms major clinical study and controlled study are located on the periphery of the map, while ergonomics is positioned toward the centre of the MCA plot.

**Figure 10:**
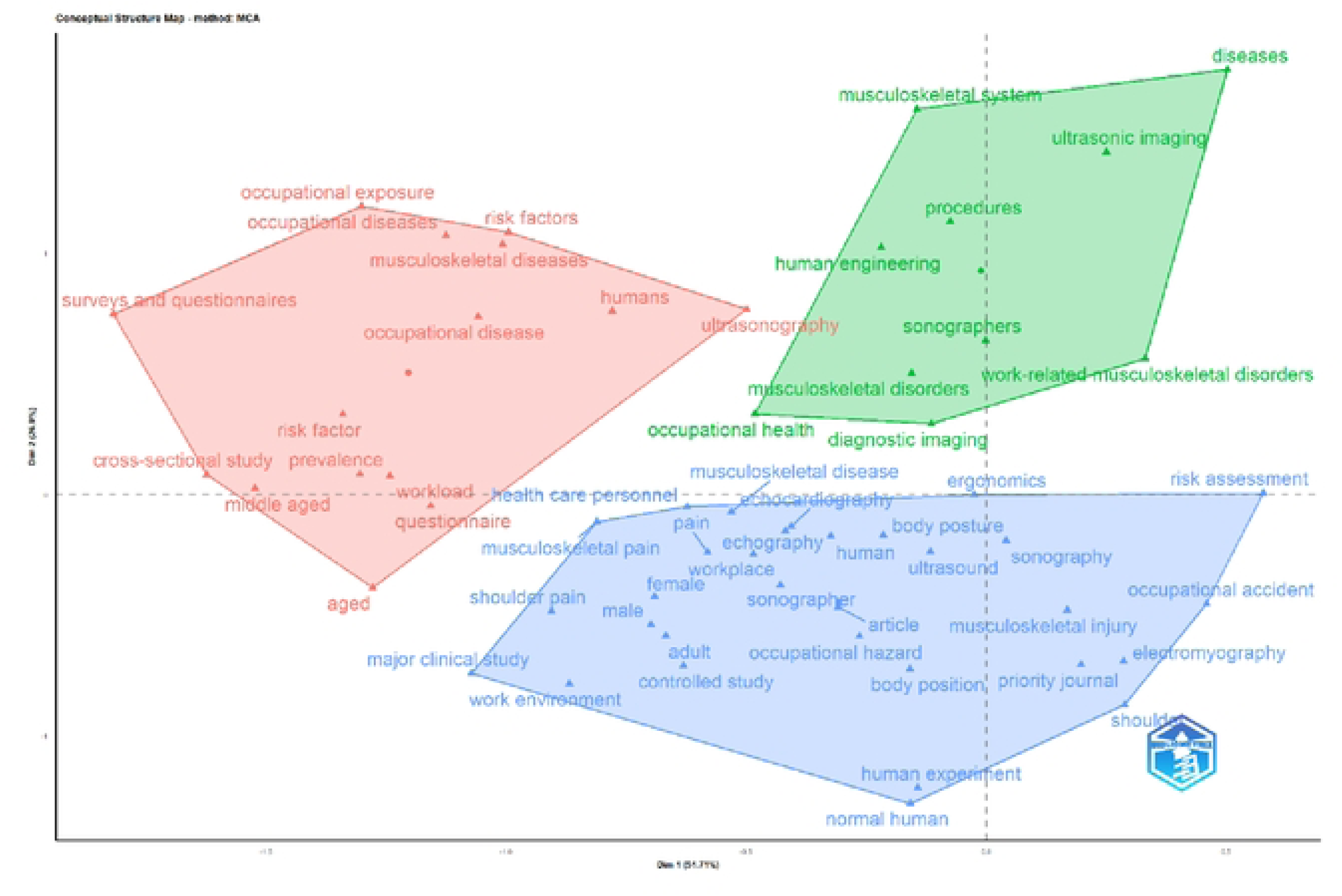
Factorial Analysis (Multiple Correspondence Analysis)

### 3.7 Most Productive Authors, Author Productivity, and Co-authorship Network

The productivity data in Figure 11 identify Evans K. D. as the author with the highest output (10 articles), followed by Forzoni L. and Sommerich C. M. (8 articles each) and Roll S. C. (7 articles). On the fractionalised article measure, Coffin C. T. has the highest fractionalised score of 3.83 from 5 articles, while Andreoni G. (4 articles, 0.73 fractionalised) and Joines S. (4 articles, 0.65 fractionalised) record lower fractionalised scores. Baker J. P., Lavender S. A., and Sanders E. contributed 4 articles each.

**Figure 11:**
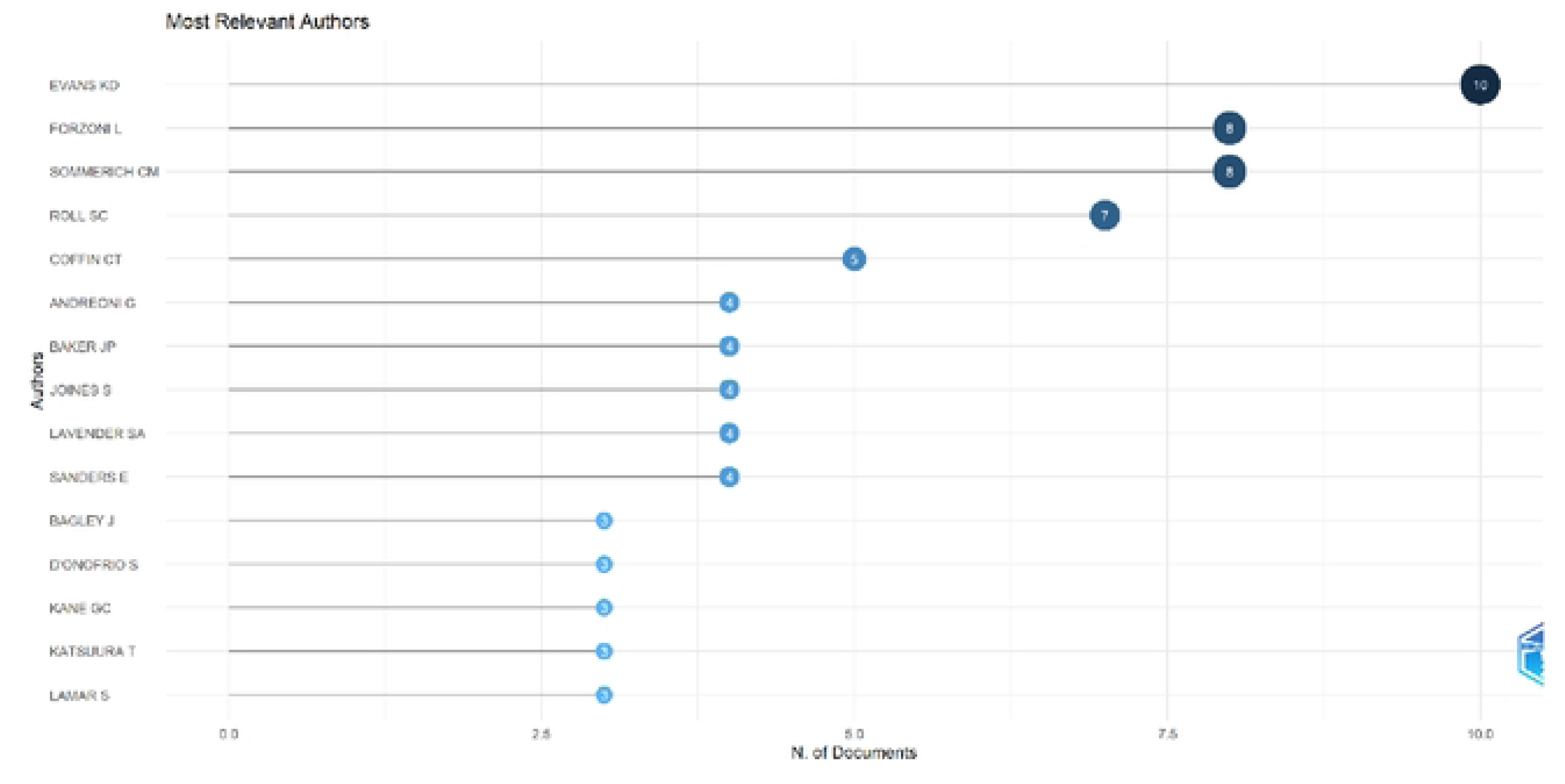
Top 15 most productive authors in sonographer ergonomics research.

Author productivity was assessed against Lotka’s law, the inverse square law of scientific productivity in which the number of authors (y) publishing (x) papers is inversely proportional to x^n^, represented by the formula x^n^ y = C. As Figure 12 indicates, 247 authors (77.2%) wrote a single document in the field, compared with the theoretical Lotka prediction of 66.2%. A small elite of approximately 1.2% of authors wrote 7 or more articles, and only 0.3% wrote 10 or more documents.

**Figure 12:**
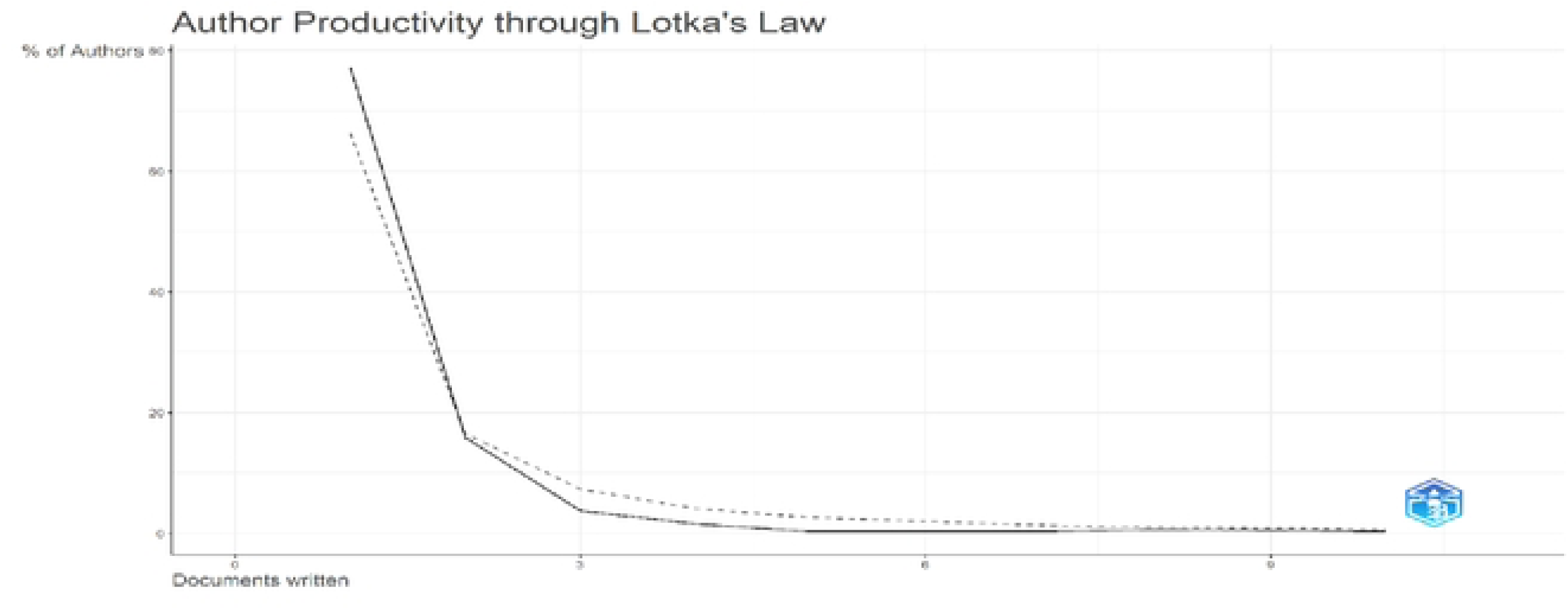
Author productivity through Lotka’s law.

The co-authorship network was built using the full counting method. Among the 322 authors identified in the 93 documents, 63 authors met the threshold of one or more documents within a linked network, as indicated in Figure 13, and were sorted into five clusters. Cluster 1 (Red, 22 items) includes Barahona O. J., Collins D. E., and Wolfman D., each with 1 document, 17 citations, and a Total Link Strength (TLS) of 23, with the same measures repeated across the cluster. Cluster 2 (Green, 14 items) contains authors with high total link strengths, including Lavender S. A. (TLS = 31), Sanders E. B. N. (TLS = 31), and Joines S. M. B. (TLS = 28), with up to 4 documents per individual. Cluster 3 (Blue, 11 items) contains Evans K. D. (10 documents, 255 citations, TLS = 49), Sommerich C. M. (8 documents, 69 citations, TLS = 44), and Roll S. C. (7 documents, 220 citations, TLS = 40). Cluster 4 (Yellow, 10 items) includes Baker J. P. (237 citations, TLS = 10) and Coffin C. T. (146 citations, TLS = 3). Cluster 5 (Purple, 6 items) centres on Bagley J. E. (3 documents, TLS = 29).

**Figure 13:**
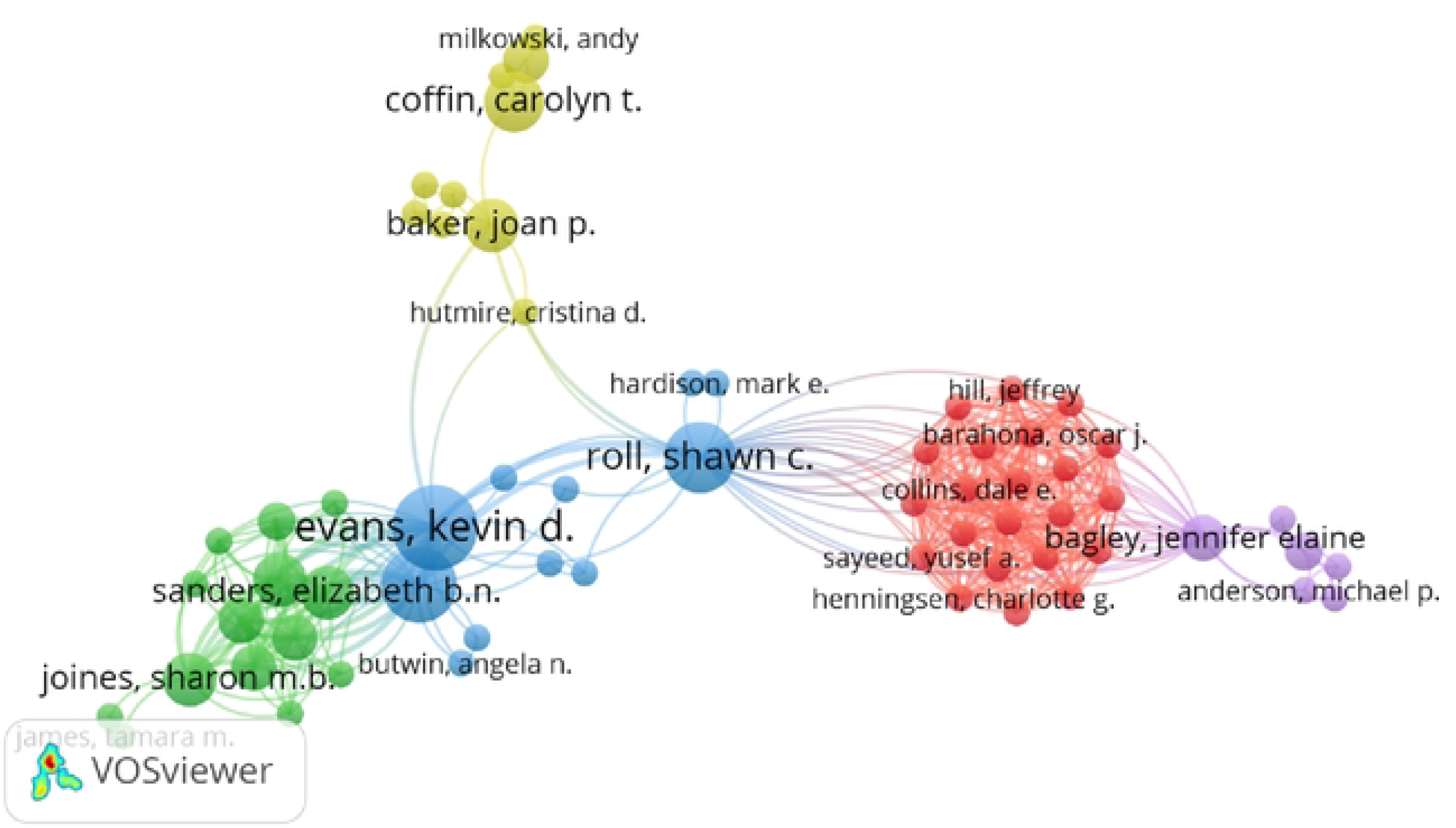
Co-authorship network among researchers.

### 3.8 Most Productive Institutions and Sources for Research Dissemination

The distribution of scholarly output across institutions is shown in Table 3. The Ohio State University leads the global ranking through its College of Engineering, Columbus (n = 8), the Ohio State University College of Medicine (n = 8), and the general body of the university (n = 5), complemented by the Mayo Clinic (n = 6) and the University of Southern California (n = 5). The Silesian University of Technology (Poland, n = 4) is the leading European centre, while Chiba University (Japan, n = 3), CQUniversity Australia (n = 3), and the University of South Australia (n = 3) represent the Asia-Pacific region. Liaquat University of Medical and Health Sciences (Pakistan, n = 3) and Imam Abdulrahman Bin Faisal University (Saudi Arabia, n = 3) also feature, and one equipment manufacturer, GE Healthcare United States (n = 2), appears among the 25 most productive institutions.

**Table 3:** Top 25 most productive institutions.

| **Affiliation** | **Country** | **Articles** |
| --- | --- | --- |
| College of Engineering Columbus | United States | 8 |
| The Ohio State University College of Medicine | United States | 8 |
| Mayo Clinic | United States | 6 |
| The Ohio State University | United States | 5 |
| University of Southern California | United States | 5 |
| Silesian University of Technology | Poland | 4 |
| Chiba University | Japan | 3 |
| CQUniversity Australia | Australia | 3 |
| Grand Valley State University | United States | 3 |
| Imam Abdulrahman Bin Faisal University | Saudi Arabia | 3 |
| Liaquat University of Medical and Health Sciences | Pakistan | 3 |
| Politecnico Di Milano | Italy | 3 |
| Seattle University | United States | 3 |
| Shahid Sadoughi University of Medical Sciences | Iran | 3 |
| University of South Australia | Australia | 3 |
| Universitätsmedizin Mainz | Germany | 3 |
| Anatomie Der Universität Basel | Switzerland | 2 |
| Arizona School Of Health Sciences | United States | 2 |
| Charles University | Czech Republic | 2 |
| Clínica Alemana | Chile | 2 |
| Diagnostic Center of the Pubblica Assistenza Di Signa | Italy | 2 |
| GE Healthcare United States | United States | 2 |
| Goethe-Universität Frankfurt Am Main | Germany | 2 |
| Guangdong Province Hospital for Occupational Disease<br>Prevention and Treatment | China | 2 |
| Hospital San José Tecsalud | Mexico | 2 |

The 93 documents were published across 44 distinct sources. As indicated in Table 4, the Journal of Diagnostic Medical Sonography (JDMS) is the clear leader, providing 21 articles, almost a quarter of the entire output. The Journal of Medical Ultrasonics (Singapore) and Work: A Journal of Prevention, Assessment and Rehabilitation contributed 4 articles each. The remainder of the top ten comprises Ergonomics (n = 3), the International Journal of Industrial Ergonomics (n = 3), the Proceedings of the Human Factors and Ergonomics Society (n = 3), the Journal of Ultrasound in Medicine (n = 3), Sonography (n = 3), Advances in Intelligent Systems and Computing (n = 3), and Procedia Manufacturing (n = 3).

**Table 4:** 10 most relevant journals.

| **Rank** | **Sources** | **Articles** |
| --- | --- | --- |
| 1 | Journal of Diagnostic Medical Sonography | 21 |
| 2 | Journal of Medical Ultrasonics (Singapore) | 4 |
| 3 | Work | 4 |
| 4 | Advances in Intelligent Systems and Computing | 3 |
| 5 | Ergonomics | 3 |
| 6 | International Journal of Industrial Ergonomics | 3 |
| 7 | Journal of Ultrasound in Medicine | 3 |
| 8 | Procedia Manufacturing | 3 |
| 9 | Proceedings of the Human Factors and Ergonomics Society | 3 |
| 10 | Sonography | 3 |

### 3.9 Top-Cited Publications

Table 5 lists the 25 most cited works in the dataset. The most frequently cited work is [8] with 147 total citations, followed by [1] with 112 citations and the highest citations per year (56.00) and normalised citation score (19.64) in the dataset. [7] recorded 97 citations, [21] 96 citations, and [24], [18], [25], and [11] 95 citations each. The list spans large-scale cross-sectional prevalence studies, biomechanical and electromyographic investigations, sub-speciality comparisons, intervention studies, practice guidelines, and systematic reviews.

**Table 5:**
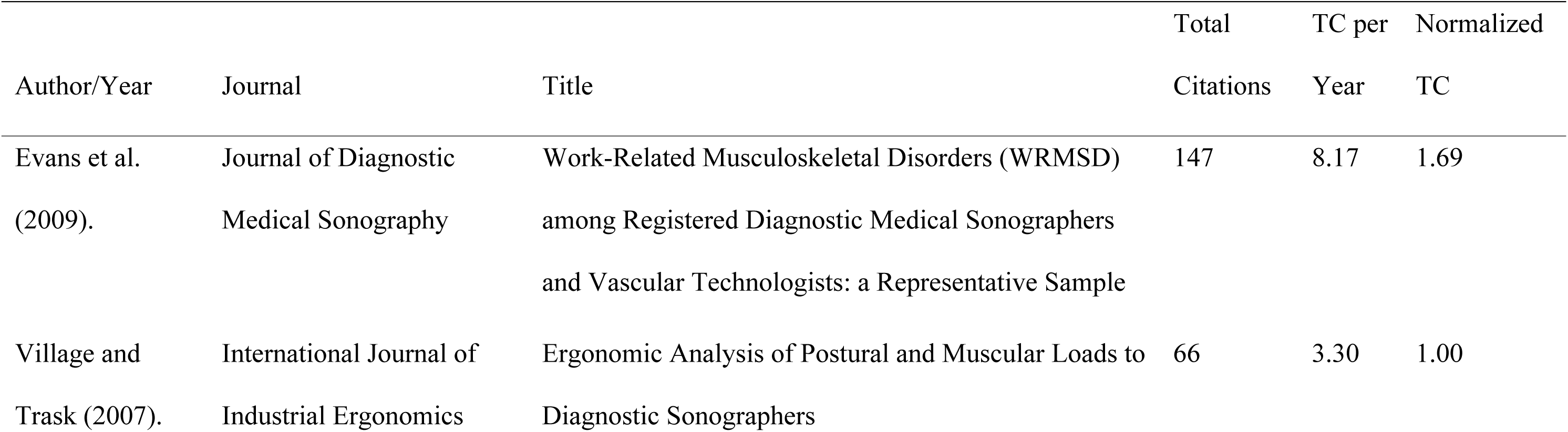
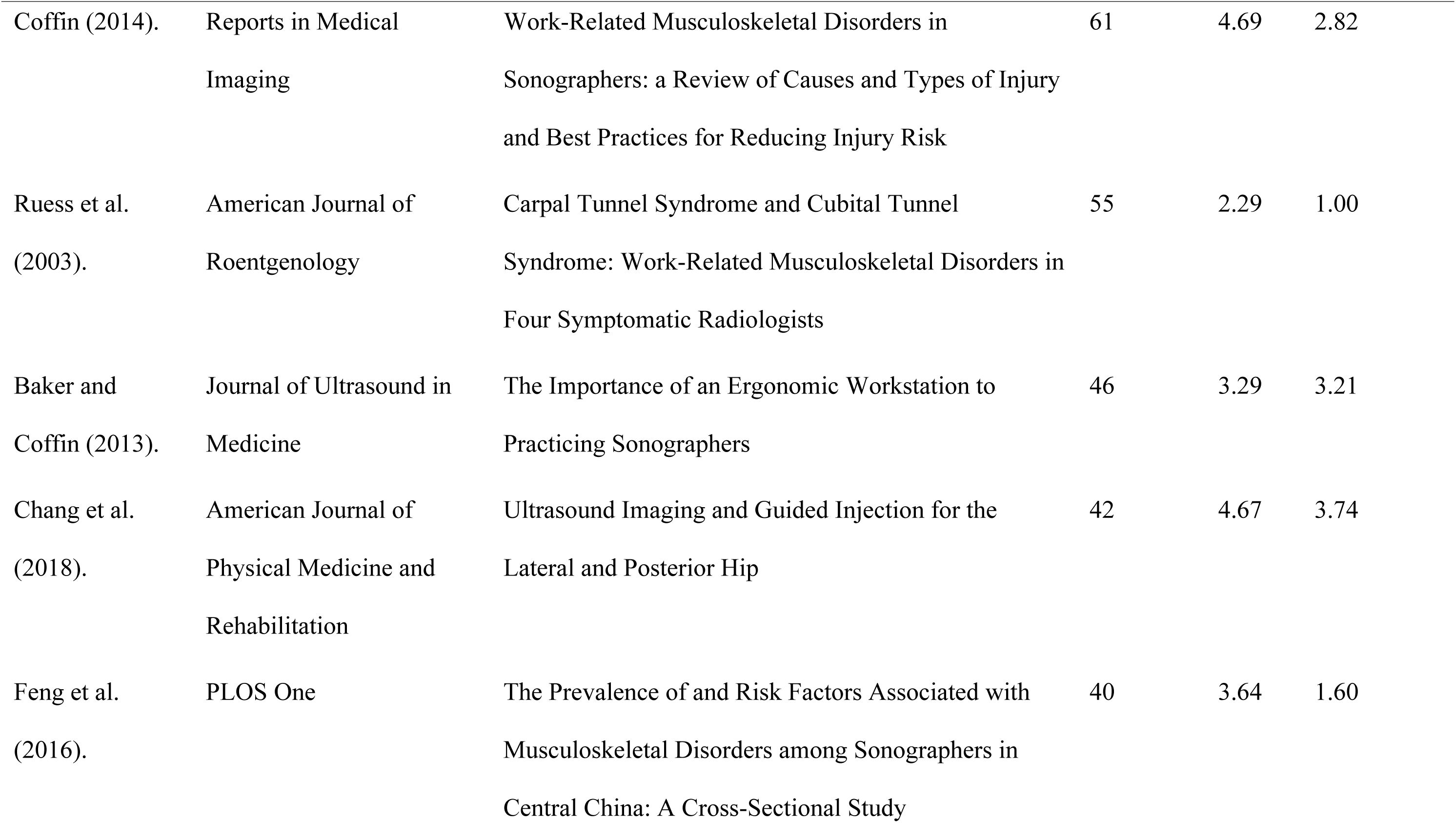
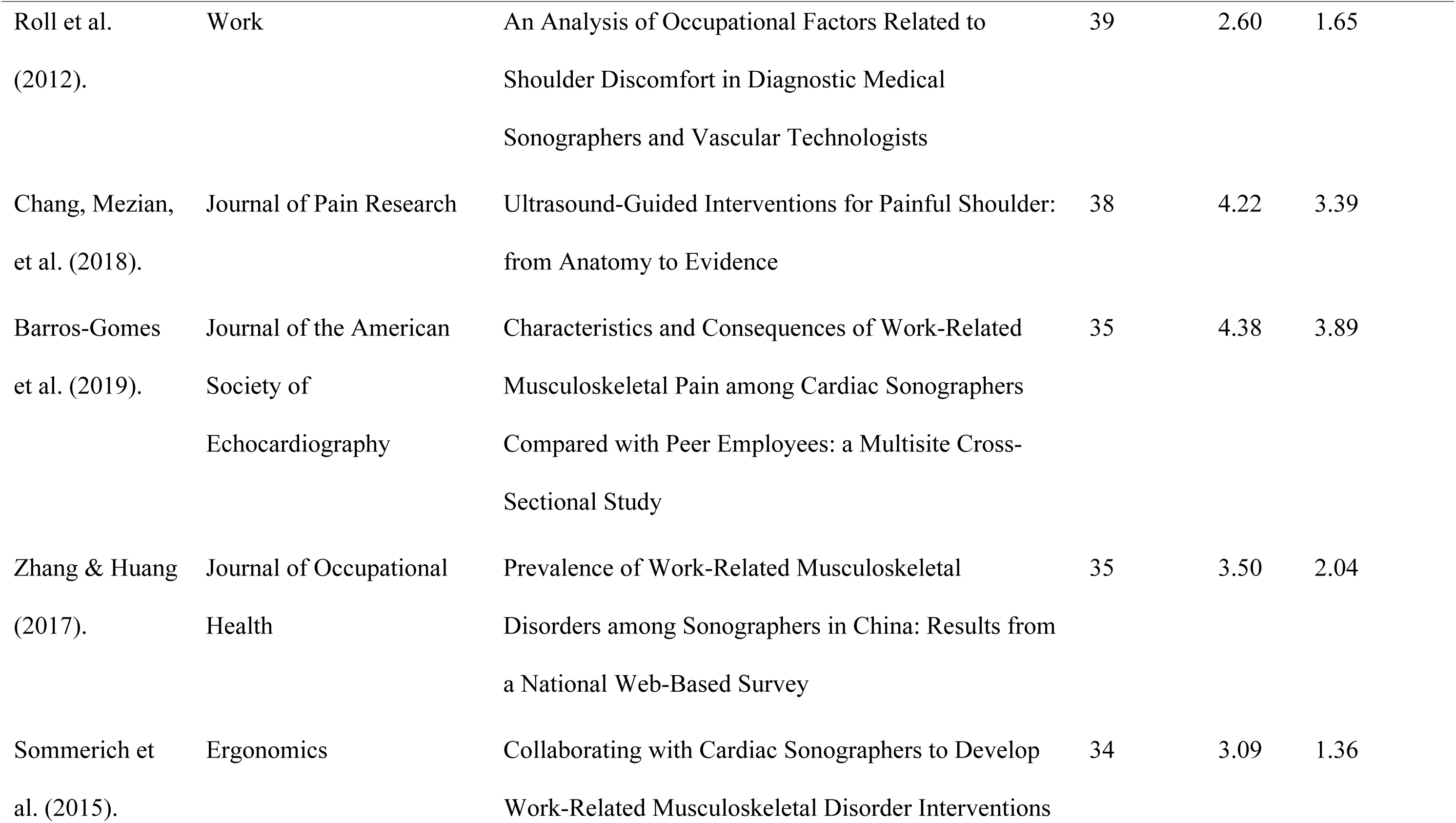
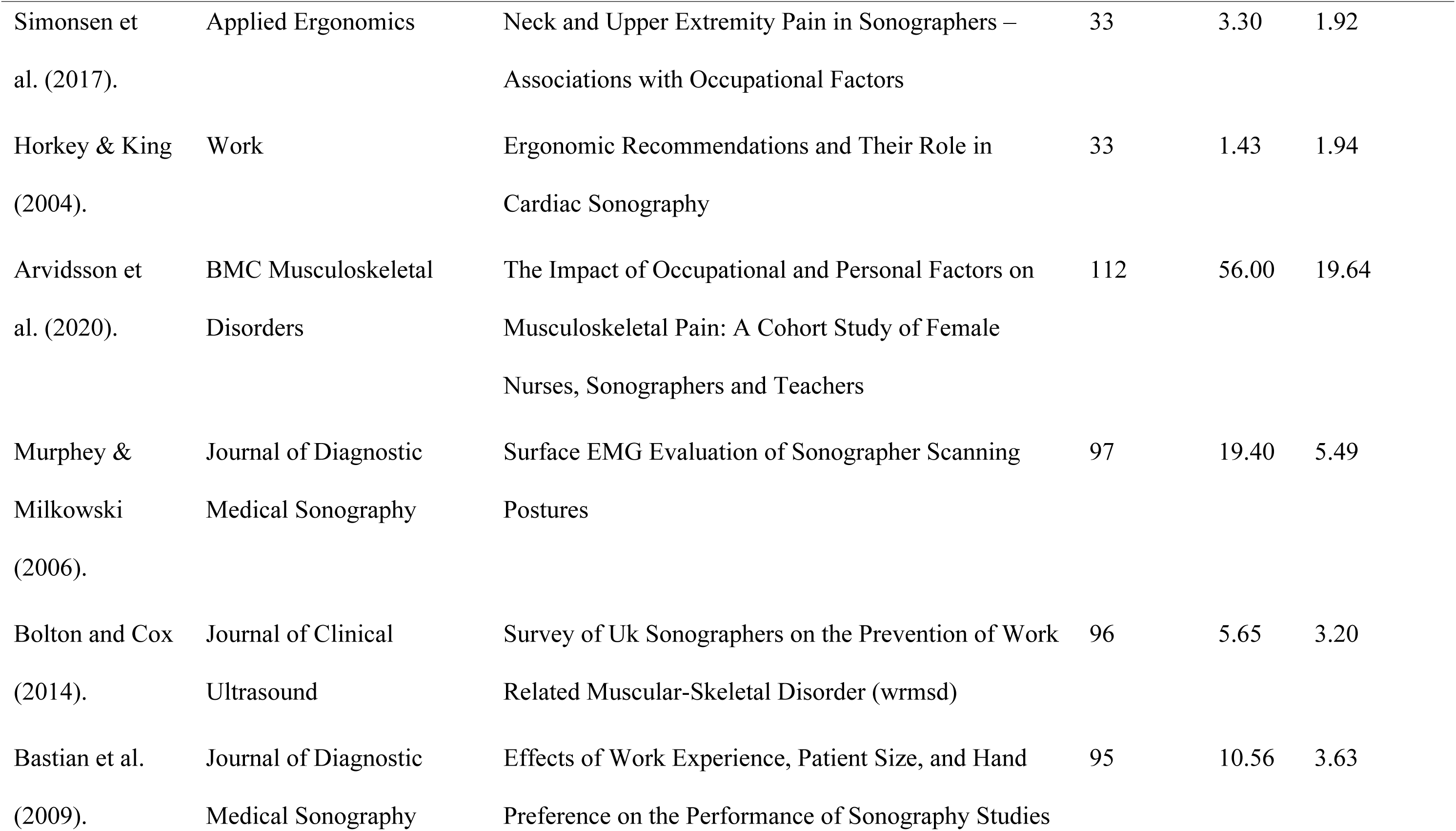
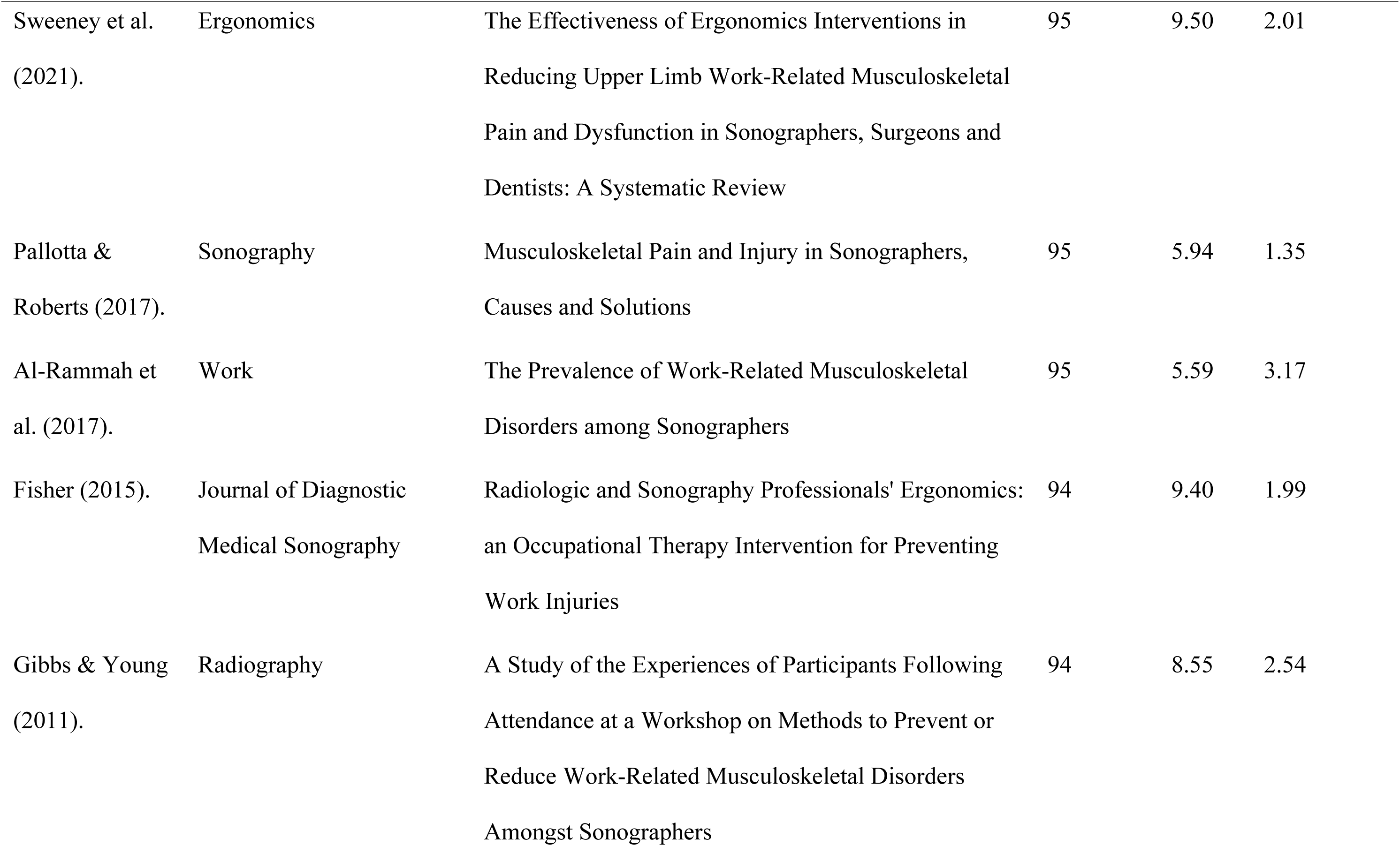
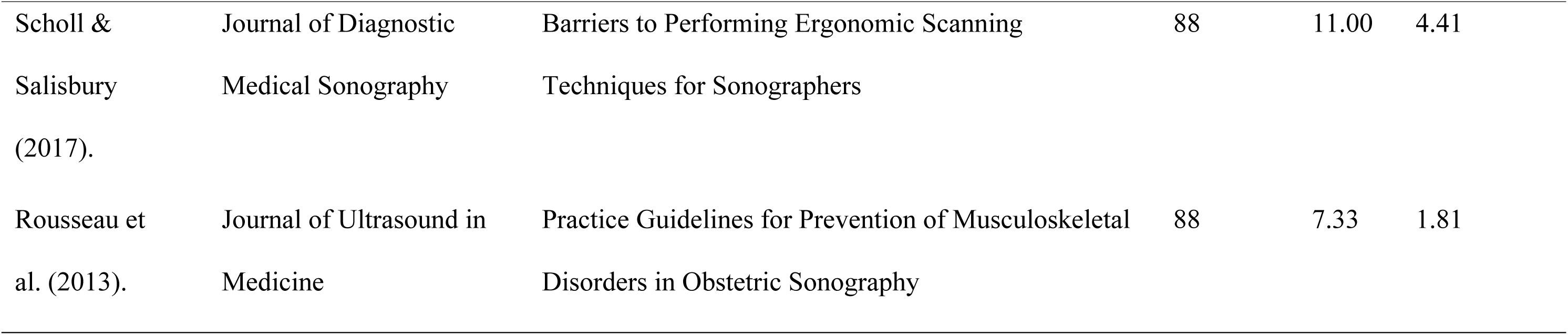
Top 25 most relevantly cited papers in sonographer ergonomics research.

## 4. DISCUSSION

### 4.1 Growth Trajectory of the Field and Its Sensitivity to External Shocks

The constant average growth rate of 8.49% per year reported in Section 3.1 indicates progressive scholarly interest in reducing work-related musculoskeletal disorders (WMSDs) through technological and ergonomic measures. The average of 14.15 citations per document indicates a strong intellectual influence, revealing that studies in this field are produced and used to guide other clinical and ergonomic investigations [27]. The dominance of original articles and reviews in the document structure demonstrates a field in which the discussion has shifted from a preliminary, conference-driven stage to more involved longitudinal studies and systematic syntheses that require peer review. The lexical richness of the discipline, evidenced by 859 Keywords Plus and 207 Author Keywords, hints at a multidimensional and interdisciplinary approach that encompasses clinical medicine, engineering, and human factors design.

The three production phases identified in Section 3.2 can be interpreted as distinct stages of maturation. The embryonic phase (2003 to 2012) indicates that the dangers of musculoskeletal disorders in sonography were known but had not yet been placed at the core of the larger occupational health debate. The surge from 2013 coincides with the global adoption of digital health norms and increased attention to the safety of healthcare workers, marking the transition to the intensification period. The 2017 to 2019 peak may indicate a golden era of ergonomic research, when adjustable workstation technologies were introduced, and high-fidelity transducer designs required empirical validation [27].

The slump from 2020 to 2023 can be explained by the global COVID-19 pandemic. The shift of healthcare research priorities to the management of infectious diseases, coupled with the excessive clinical workload imposed on sonographers during the crisis, suffocated ergonomic field studies and longitudinal interventions in the short run. This observation highlights how prone niche occupational health research is to external systemic shocks [26,29]. The resurgence recorded in 2024 and 2025 reflects a new academic interest potentially triggered by the advent of Artificial Intelligence (AI) and robotic-assisted ultrasound, which signal a potential paradigm shift in the ergonomic stress of the profession. Although the growth rate of 8.49% is not as high as the 15.48% reported in larger studies on digital governance [26,33], it indicates the emergence of a mature and stable field. The trend highlights that as sonography technology advances, the scientific community is responding with a commensurate focus on the physical and cognitive well-being of practitioners.

### 4.2 Geographic Concentration and the Global Equity Gap

The social network of a research area is determined by the intensity and geographic scale of its scientific partnerships, which are proxies for the movement of intellectual capital and technological know-how. The strong USA-China and Italy-China linkages reported in Section 3.3 imply a triadic interaction of knowledge between North America, Europe, and East Asia, presumably focused on the co-development and biomechanical trials of ultrasound equipment and ergonomic software interfaces [26,27]. The cohesive European sub-network of Italy, Germany, and Switzerland probably reflects similar standards of occupational health and medical safety across Europe. The inter-regional links between Sweden and Bahrain, the USA and Iran, and the USA and Saudi Arabia demonstrate that the ergonomic issues linked with medical imaging are treated as a global health issue requiring transcontinental research that goes beyond conventional geopolitical ranges to understand the commonality of sonographer strain [28].

The international co-authorship rate of 9.68%, however, implies that sonographer ergonomics research is still conducted in national or regional clusters rather than in a large-scale international network. This result is consistent with past evidence in occupational health bibliometrics, which has presented interventions as customised to particular national healthcare systems or local institutional safety standards [28]. A critical examination of the network also shows a high level of geographic centralisation. While the USA and China serve as the main channels of research, collaboration with South America and other emerging economies is very limited, and countries such as Chile and Mexico occupy peripheral positions decoupled from international research activities. This bias reflects larger occupational health bibliometric patterns in which the narrative of intervention technologies is dominated by high-resource settings. It poses a threat that ergonomic solutions, mostly developed and tested in Western clinical contexts, leave sonographers in low-resource settings, who may be using older and less adjustable equipment, as thematic outliers of the global safety discussion. The institutional data in Section 3.8 reinforce this concern: the dominance of the Ohio State University hub, alongside the Mayo Clinic, represents a developed interdisciplinary paradigm in which bioengineering concepts are directly translated into the clinical issues of the sonography workforce, and implies that modern global standards of sonographer safety are largely influenced by North American biomechanical and clinical research standards. The presence of GE Healthcare among the 25 most productive institutions underscores an unusual industry-academic overlap and the necessity of synergy between empirical studies and the commercialisation of ultrasound equipment, indicating that institutional productivity in this line is not just an academic exercise but a practical endeavour aimed at redesigning the physical tools of the profession.

### 4.3 Conceptual Structure: From Hazard Identification to Technology-Led Prevention

The keyword profile in Section 3.4 captures a niche, clinically based, and intensely focused research front concerned with the physiological integrity of the human operator in the sonographic setting. The prominence of human and ergonomics establishes the core unit of analysis and the main disciplinary reference point, relating the physical requirements of sonography to occupational safety and equipment design. The frequency of echography, musculoskeletal disease, and ultrasonography indicates a well-developed area in which the physical cost of medical imaging is meticulously reported in the context of diagnostic technology, while the high rank of the keyword article indicates a strong presence of original empirical research over theoretical or editorial contributions. The frequency of adults confirms that the research is focused on the professional workforce rather than paediatric or academic populations. The near parity of females and males suggests a focused effort within the scientific community to address the gender-specific nuances of musculoskeletal strain. In a profession whose workforce tends to be gender-specific in both demographic and anthropometric terms, this signals that ergonomic interventions are becoming highly specific rather than generic.

The trend topic sequence reported in Section 3.4 depicts the conceptual path of the field over two decades: a predominantly reactive early period concentrated on identifying the mechanical causes of strain and developing a bioengineering platform of intervention; an intermediate period of quantitative empirical and methodological maturation in which passive observation gave way to active human experiments and isolated physical complaints were given the wider framework of clinical diagnoses and occupational health; and a current period oriented toward large-scale validation and symptomatic distress management. The persistence of musculoskeletal pain and the questionnaire as high-frequency terms indicates that, twenty years on, the subjective experience of pain remains an important indicator of occupational failure, and that the field is no longer a matter of identifying basic posture but of longitudinal, systemic intervention to curb the endemic nature of injury.

The cluster analysis in Section 3.5 reveals a high level of intellectual hierarchy. Cluster 1 functions as the empirical engine of the field, concerned with the mechanical relationship between the practitioner and the workstation and representing its best peer-reviewed and synthesised component. Cluster 2 denotes the institutional framework of the field, in which clinical practice terms are connected to legal and safety terms; the inclusion of the keyword United States of America in this cluster demonstrates the relevance of North American regulatory and academic organisations in the formal classification of WMSDs. Cluster 3 captures the subjective experience of the workforce and how institutional factors, including workload and ageing demographics, contribute to the shift from minor strain to chronic pain. The critical contrast is that Cluster 1 emphasises the act of scanning while Cluster 2 emphasises the consequences and the labels applied to them systemically. Biomechanical data are strongly coupled with pathological labels, yet there is an evident disparity in how such objective measures inform the workload problems found in Cluster 3. While the literature quantifies muscle contraction and the nosology of the resulting disorder, the mapping indicates a relative lack of interaction in developing interventions that directly reduce the pain reported in epidemiological surveys. Future studies should merge the biomechanical rigour of Cluster 1 with the workload realities of Cluster 3 to produce more holistic ergonomic solutions.

The thematic map in Section 3.6 exposes a parallel surveillance-intervention gap. The status of major clinical study and risk factor as Motor Themes suggests a change from theoretical ergonomics toward rigorous evidence-based clinical surveillance, bridging the subjective experience of healthcare personnel with proven epidemiological measures. However, the location of demographic-specific biomechanical testing (electromyography, human experimentation) in the Niche quadrant indicates that these interventions, though internally strong, have yet to be incorporated into mainstream hospital safety practice. The Basic Themes quadrant confirms that terms such as occupational disease and human engineering tend to serve as background descriptions rather than developing fields of fresh research, while the position of occupational accident and musculoskeletal injury among declining themes signals a conceptual shift from acute injury control to chronic, cumulative WMSD prevention. Although the scientific community is now very competent in recording the extent of the problem, the technical solutions, including bio-engineered workstation layouts and real-time biofeedback, remain in silos. To advance the field, a strategic shift is needed to move these Niche technological interventions onto the Motor agenda so that advanced bioengineering becomes an integrated part of clinical practice.

The thematic evolution reported in Section 3.6 shows a definite conceptual narrowing followed by a systemic widening. Between 2003 and 2021, the research narrowed from broad occupational hazards to very specific sonographer-focused WMSDs, and from 2022, this particular knowledge has been re-contextualised within larger occupational health frameworks. Ergonomics has served as the transversal theme, the stable bridge, throughout all periods, while the keywords around it have developed from reactive (hazard, injury) to proactive (awareness, exercise, health policy). Nevertheless, high-frequency keywords associated with pain and disease persist even in the most recent slice. This opposition implies that, although the scientific interpretation of ergonomics has become a more advanced, policy-focused dialogue, the clinical reality of the sonographer is still characterised by a high level of musculoskeletal distress, and a gap remains between research and efficient application at the workplace. The factorial analysis reinforces this reading: the separation of the clinical-experimental cluster from the epidemiological-diagnostic cluster in the MCA space points to a conceptual gap between the two measurement traditions, while the central position of ergonomics shows that it is the unifying variable connecting clinical practice, disease classification, and human experiments. The peripheral isolation of major clinical studies and controlled studies suggests that high-level evidence-based interventions remain a professionalised subgroup rather than the normative centre of the ergonomic discourse. Present studies ought to focus on centralising controlled clinical interventions to close the gap between experimental biomechanics and population-level prevalence.

### 4.4 Intellectual Leadership and the Concentration of Knowledge

The productivity profile in Section 3.7 shows that a small and specialised community of researchers occupies the intellectual field [27]. The significant contributions of Evans, Forzoni, Sommerich, and Roll show a long-term dedication to exploring the longitudinal effects of sonographic work. The fractionalised scores add nuance: Coffin C. T. is frequently the main or sole investigator of their works, indicating a high level of personal intellectual leadership, whereas authors such as Andreoni G. and Joines S. appear to work in larger, multi-disciplinary research groups. This differentiation is essential: it implies that, though the area benefits from large-scale collaborative biomechanical research, it is also highly influenced by committed individual specialists who provide the theoretical and clinical foundations of ergonomic standards. The presence of a maturing secondary level of researchers points to a productivity distribution following a Price-like law in which a small percentage of authors contribute a large proportion of high-impact publications. This concentration of the knowledge base among a handful of anchor authors implies that the methodological preferences of these authors, such as workstation design and scanning technique, are highly influential in shaping worldwide standards of ergonomic best practice.

The Lotka distribution fits a specialised medical niche in which a large number of clinical practitioners contribute a single case study or local intervention without sustaining a long-term research programme in ergonomics. The prolific core, in which scholars such as Evans and Forzoni are included, drives the theoretical and methodological standardisation of the field, and the fact that only 0.3% of authors wrote 10 or more documents indicates that ergonomics knowledge in sonography is not universally distributed through the medical community but is held by a few research centres and educational institutions. A critical inference from this distribution is that there is a high barrier to entry for sustained ergonomic research. The large share of one-time contributors (77.2%) indicates that although many scholars acknowledge the significance of ergonomics, not all are supported by institutions or longitudinal data to move beyond exploratory research. The concentration of influence within the core guarantees uniformity in ergonomic recommendations but poses a risk of intellectual silos. To advance the field, mentorship programmes and collaborative networks are needed to convert occasional authors into prolific ones, which will diversify the geographic and clinical views in the sonographer safety debate.

The co-authorship network supports the same conclusion. Cluster 1 appears to represent a high-impact, multi-institutional position paper or white paper focused on defining professional standards, functioning as the regulatory core of the field. Cluster 2 operates as a technical laboratory bridging human factors engineering and clinical sonography. Cluster 3 is the intellectual and prolific centre whose members act as key bridges ensuring knowledge transfer between experimental biomechanics and clinical sonographer safety. Cluster 4 displays the professional advocacy and clinical standards group: although its members have high citation impact, their relatively low link strengths imply that their foundational contributions are often the product of independent clinical practice or smaller professional groups rather than large corporate networks. Cluster 5 points to specialised nodes of research with strong internal connections. Nevertheless, the reality that only 63 of 322 authors belong to a major connected network is indicative of the still prevalent isolated pockets in which sonographer ergonomics research is conducted. Such disjointed cooperation means that, though the field is well led, further interaction across the clusters is needed to ensure that biomechanical evidence and professional guidelines are more frequently combined into the standard clinical body of literature. The source analysis in Section 3.8 adds a dissemination dimension: the pre-eminence of the Journal of Diagnostic Medical Sonography establishes it as the main venue for peer-reviewed research on the professional health of the sonography workforce, while the presence of Work and the human factors and engineering titles shows that clinical ultrasound journals record injury prevalence while specialised human factors and engineering platforms are used to propose and test technical solutions, including the application of automation and manufacturing design to alleviate sonographer strain.

### 4.5 From Awareness to Environmental Transformation: Evidence from the Most-Cited Works

Citation analysis serves as a proxy for scientific impact, identifying the intellectual pillars that have historically defined the risks, prevalence, and interventions associated with WRMSDs. Large-scale cross-sectional studies form the basis of the literature, setting a substantial international incidence of WRMSDs. The most frequently cited work, [8], served as a critical benchmark, stating that 90% of sonographers scan while in pain. This finding is consistent with other previous studies [12,14]. More recent data from [9] and [20] indicate that in more densely populated regions such as China, prevalence rates approach 100%. While this trend reflects improved awareness, it also suggests that traditional educational initiatives have reached their limit, failing to provide further measurable reductions in injury rates. Although [23] demonstrated that multimedia tutorials can enhance awareness and practice, the high incidence rates reported by [8] suggest that system demands tend to override behavioural change.

These high prevalence rates are rooted in physiological stressors documented by the biomechanical evidence of high-impact studies. [13] using video-based postural analysis, revealed that sonographers dedicate 68% of their scanning period to dangerous shoulder abduction (greater than 30 degrees). This is empirically confirmed by the surface electromyography study [7], who demonstrated that reducing abduction from 75 degrees to 30 degrees can result in a 50% decrease in muscle activity, with a further reduction in load of 88% through forearm support. Alongside this physiological evidence, [10] outlined a set of obstacles to the adoption of these optimal postures, namely patient obesity, the volume of portable examinations, and time constraints. This poses a fundamental conflict within the literature: [24] indicated that the risks of large patient habitus can be reduced through ambidextrous scanning and greater skill, whereas [10] held that the majority of these issues are beyond the control of the sonographer.

A comparison of sub-specialities reveals that certain diagnostic roles impose a greater ergonomic burden than others. [6] noted that 86% of cardiac sonographers reported musculoskeletal pain with a higher impact on labour activities compared with 46% in the control group, and [5] reported that 80% of obstetric sonographers experienced pain and suffered worsening visual acuity. These results advocate a change from a one-size-fits-all approach to specialised intervention. [17] advanced this concept by suggesting a participatory approach in which engineers and ergonomists work closely with cardiac sonographers to develop technical prototypes. This shift toward participatory ergonomics represents a philosophical change away from imposed mandates and toward user-driven solutions.

The psychosocial and longitudinal dimension of the profession is a crucial focus of contemporary research. [1] showed that psychosocial factors and previous pain history are the best predictors of future injury, suggesting that the ergonomic crisis is not only the result of physical strain but also of workplace stress and job dissatisfaction [19]. [25] cautioned that failure to intervene could lead to career-ending injuries in a third of all sonographers, and the regional study by [11] in Saudi Arabia directly correlated the severity of pain with daily patient throughput and cumulative years of service.

Taken together, these 25 articles indicate a discipline at a crossroads. Although the biomechanical effects of sonography are well characterised through electromyography and postural measurement [7,13], and clinical outcomes such as carpal tunnel syndrome were reported as early as [3], the remedies remain unclear. The systematic review by [18] identified a significant gap in ergonomic efficacy, noting that only microbreaks and specialised transducer handle designs are supported by moderate to strong evidence for pain reduction, while most traditional interventions lack high-quality empirical support. This issue is heightened by the decay of continuous professional development: [22] and [21] observed that the benefits of ergonomic workshops and academic training typically diminish within three months without constant reinforcement. Consequently, the challenge of sonographer health has shifted: it is no longer merely about identifying risks but about initiating fundamental environmental and organisational transformation. [15] and [2] [identified a significant paradox in which ultrasound, a vital tool for diagnosing musculoskeletal disorders, remains a key contributor to the musculoskeletal disorders affecting the sonography workforce at an unprecedented rate.

### 4.6 Limitations

This study has some limitations. The use of the Scopus database may have omitted research in specialised regional journals that are not globally indexed. In addition, the emphasis on peer-reviewed articles excludes grey literature, including industry-based ergonomics reports, safety policy notes released by professional bodies, and clinical guidelines, which contain valuable information on sonographer health. Lastly, the 2025 publications are partial-year figures that were not treated as finalised annual figures but were interpreted descriptively.

## 5. CONCLUSIONS AND WAY FORWARD

The research is the first systematic review and bibliometric analysis of the literature on ergonomic intervention and technological aids to prevent work-related musculoskeletal disorders (WRMSDs) in sonographers from 2000 to 2025. Combining the PRISMA framework and automated metadata analysis, the study was able to map the intellectual structure, thematic development, and collaborative networks of an area of research that is important to the sustainability of the diagnostic imaging workforce. The results show that the interest among scholars has been on the rise, but the professional reality is still marked by an unminimised occurrence of injury, which is predetermined by the necessity to make a radical change in the approach towards the management of the ergonomic risks in the clinical setting.

Quantitative milestones that the research had determined highlight a long-standing and growing crisis. Examination of the curated data indicates that regardless of decades of educational interventions, the sonographer prevalence of pain has remained approximately at 90%, and regional data indicate that the prevalence has attained up to 100%. The findings indicate that the field has passed through different stages, with early stages of documenting prevalence and clinical outcomes, including carpal tunnel syndrome, and a biomechanical phase, which employs electromyography and postural mapping to measure dangerous shoulder abduction and transducer pressure. Currently, the landscape has shifted to a technological and participatory stage with the emphasis laid on the design of specialised handles, workstation automation, and administrative controls like micro breaks.

Thematic and conceptual mapping have reflected that knowledge is no longer the main barrier to preventing injuries, but environment and organisational limitations. The review of literature with high impact has shown that ergonomics is often compromised by factors such as high prevalence of patient obesity, portable examination needs, and high-volume schedules. Consequently, the conventional approach of personalised individual posture correction is not enough. A new paradigm is required, which considers technology and organisational policy as separate, underlying instruments of sonographer safety. The collaboration mapping also reveals that there is a strong performance of international research networks, but a severe necessity for more participatory research where sonographers are directly engaged in the development and testing phase of new ergonomic devices.

To guide the future of the field, the following “Way Forward” addresses the specific research gaps and requirements identified through this bibliometric mapping, as shown in Table 6:

**Table 6:**
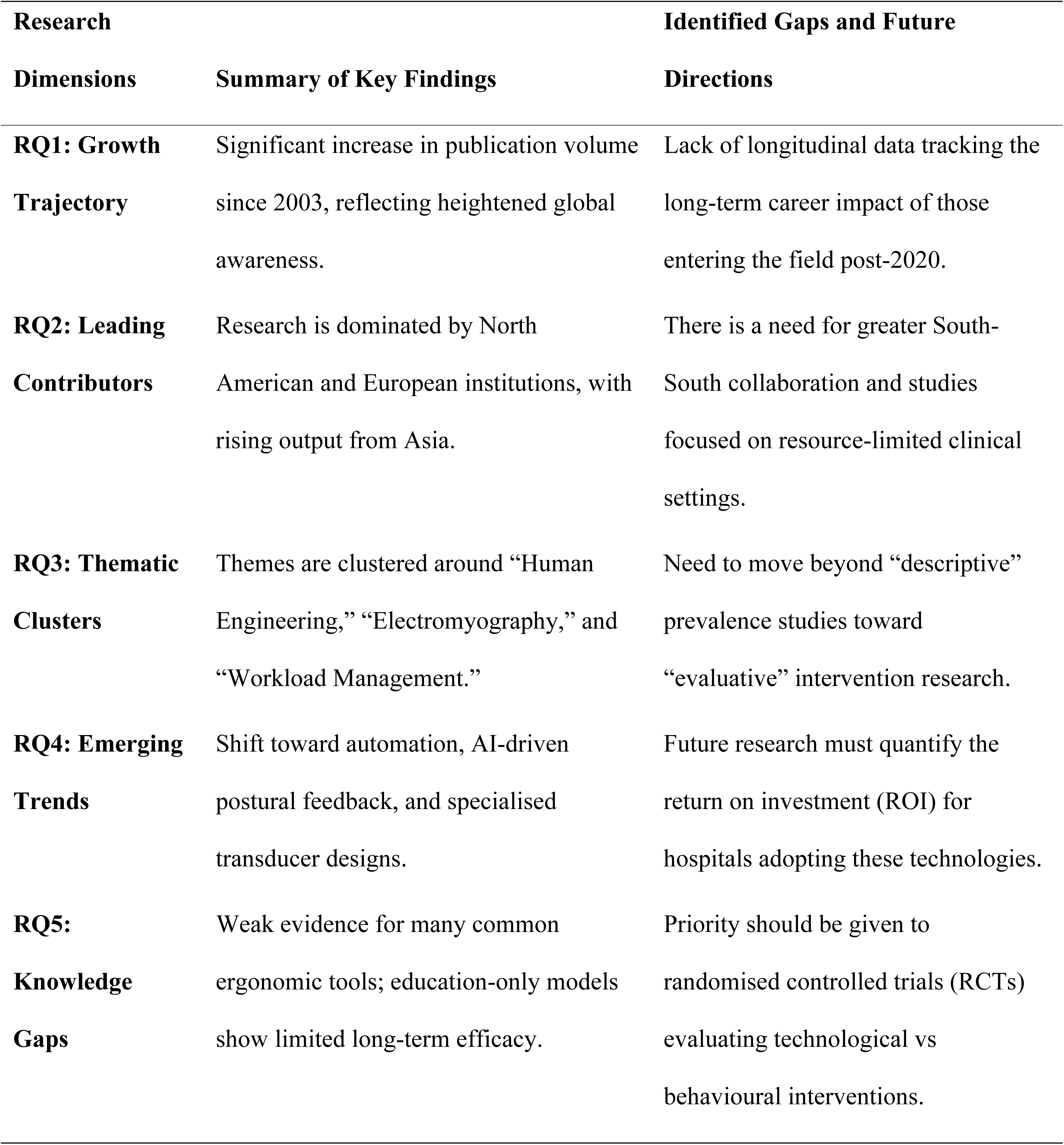
Summary of key findings and implications.

Furthermore, the future of sonographer ergonomics depends on the transition from mere awareness to tangible change in the environment. Hospital administrators and practitioners should focus on the introduction of high-order interventions, including automated workstations and mandatory microbreak software, that reduce the risk regardless of the actions of the operator. The healthcare sector can reduce the rate of professional injuries, which have witnessed little improvement in over two decades, by linking research agendas with technological innovation and organisational change. This research acts as the guide that will see that transition, making the workforce of physical and professional sustainable in the long term of the diagnostic medical sonography field.

## Data Availability

The data supporting the findings of this study are available from the corresponding author upon reasonable request

## Acknowledgement

We wish to express our sincere gratitude to Engr. Chris Kurbom Tieru for his immense assistance from the start to the completion of the manuscript.

